# The Impact of COVID-19 Lockdowns on Mental Health Patient Populations: Evidence from Medical Claims Data

**DOI:** 10.1101/2021.05.26.21257598

**Authors:** Ibtihal Ferwana, Lav R. Varshney

## Abstract

**Background:** Social distancing policies were enacted during March 2020 to limit the spread of COVID-19. Lockdowns and movement restrictions increased the potential of negative impact on population mental health, in which depression and anxiety symptoms were frequently reported by different population groups during COVID-19 lockdown. However, the causal relationship of mitigation policies on national-wide mental health resource usage is lacking.

**Objective:** This study investigates the effect of COVID-19 mitigation measures on mental health across the United States, on county and state levels. It examines the effect on mental health facility usage and the prevalence of mental illnesses on the total population, different age and gender groups, and patients of selected mental health diagnoses.

**Methods:** We used large-scale medical claims data for mental health patients dated from September 1, 2019 to December 31, 2020, with publicly available state- and county-specific COVID-19 cases from first case in January to December 31, 2020, and used publicly available lockdown dates for states and counties. We designed a difference-in-differences (DID) model, which infers the causal effect of a policy intervention by comparing pre-policy and post-policy periods in different regions. We mainly focused on two types of social distancing policies, stay-at-home and school closure orders.

**Results:** Based on common pre-treatment trend assumption of regions, we find that lockdown has significantly and causally increased the usage of mental health in regions with lockdowns in comparison to regions without. In regions with lockdown orders the resource usage increased by 18% compared to 1% decline in regions without a lockdown. Also, female populations have been exposed to a larger lockdown effect on their mental health with 24% increase in regions with lockdowns compared to 3% increase in regions without. While male mental health patients decreased by 5% in regions without lockdowns. Patients diagnosed with *panic disorders* and *reaction to severe stress* both were significantly exposed to a significant large effect of lockdowns. Also, *life management difficulty* patients doubled in regions with stay-at-home orders but increased less with school closures. Contrarily, *attention-deficit hyperactivity* patients declined in regions without stay-at-home orders. Patients older than 80 used mental health resources less in regions with lockdowns. Adults between (21 – 40) years old were exposed to the greatest lockdown effect with increase between 20% to 30% in regions with lockdown.

**Conclusion:** Although non-pharmaceutical intervention policies were effective in containing the spread of COVID-19, our results show that mitigation policies led to population-wide increase in mental health patients. Our results suggest the need for greater mental health treatment resources in the face of lockdown policies.

## 1 Introduction

As the COVID-19 pandemic began and confirmed cases rose, people voluntarily stayed at home and limited their trips in weeks before public policy interventions were imposed [34]. Subsequently, social distancing policies were issued globally as a form of non-pharmaceutical intervention, including limiting people’s gatherings, closing schools, and fully restricting movements by lockdown orders (also called stay-at-home or shelter-in-place orders) [40], so as to contain virus spread [48, 3, 3, 23]. Yet, studies indicate lockdowns affected mental health parameters more than the evolution of the pandemic itself [46]. Indeed, both personal and community responses of self-isolation and public policies raised mental health concerns among the population, with numerous calls for preventive psychological interventions [16, 13, 10, 33]. Hence, examining the causal effect of COVID-19 lockdown measures on mental health is important for health and economic planning during pandemics.

Mental disorders have been more economically costly than any other disease. In 2013, mental disorders were the leading segment of health care spending in the United States [45]. It has been estimated that half of the global economic burden will be attributable to mental illness [6]. Poor mental health might come with additional health system costs due to the increase risks of physical morbidity [36]. On an individual-level, common mental illnesses have been associated negatively with income [43], and have been related to lower levels of productivity at work [9]. Additionally, there is a huge lifetime cost to education—with increased school drop-out [11]—and criminal justice sectors due to poor mental health, which even outweighs the health care system cost [4]. Mental health has been related to social capital on individual and community levels [37, 5]. Indeed, good social capital plays a role in promoting healthier public behaviors, especially during COVID-19, social capital has been associated with higher levels of vaccination rates, self-isolation, and face masking [21]. The risk of mental health degradation goes beyond to impact the advantage of social capital in the face of viral diseases. Given these consequences of poor mental health on overwhelming the health care systems [47], it has been essential to mitigate additional mental degradation and avoid potential future economic and social costs.

There have been several observations of mental health degradation during the COVID-19 pandemic mainly through examining associations with various indirect measures [42, 51, 22]. Reported suicidal thoughts and depression symptoms almost quadrupled from 2019 to June 2020 [14] and about 30% more respondents^1^ reported depression and anxiety symptoms than in 2019 in the United States [1]. Other studies focused on the association between movement restrictions and mental health, in which reduced physical activity was found to be a leading factor to higher rates of depression during the pandemic [38, 25, 15]. In addition to self-reported and measured depression rates, mental health has also been observed online by tracing people’s keyword searches. Within the first two weeks after lockdowns, the search for words such as *worry, sadness*, and *boredom* peaked in several US states, which has been significantly associated with negative feelings [19, 7].

Although previous work shows degradation of mental health during the COVID-19 pandemic, this mainly depends on self-reported questionnaires or online behavior patterns which do not reflect the actual usage of mental health services or the rising need for mental health treatment. Examining the use of mental health resources and the prevalence of mental illnesses would further help in measuring the actual cost of COVID-19 lockdowns on mental health and inform mental health treatment resource planning for future lockdowns. To the best of our knowledge, there is no large-scale study that has investigated the effect of extended lockdown on the usage of mental health resources across the country. We empirically estimate the *causal* effect of COVID-19 distancing policies on mental health across counties and states in the United States by comparing the differences in changes between locked and non-locked down regions using a large-scale medical claims dataset that covers most hospitals in the country. We use the number of patients visiting mental health facilities as a measure for the usage of mental health resources, and we consider emergency department (ED) visits for mental health issues as a proxy for the development of new mental disease, here, so severe that treatment could not be avoided. The usage of mental health resources can further trigger analysis on economic costs borne by health care systems and the country as a whole. Mental health ED treatment visits might further reflect the mental health cost on an individual level.

Our results show that extended lockdown measures significantly increase the usage of mental health resources and ED visits. In particular, mental health resource usage in regions with lockdown orders have significantly increased by 18% compared to 1% decline in regions without a lockdown. Female populations have been exposed to a larger lockdown effect on their mental health with 24% increase in regions with lockdowns compared to 3% increase in regions without, while male mental health patients decreased by 5% in regions without lockdowns. Cases of *panic disorders* and *reaction to severe stress*, and *life management difficulty* significantly increased by lockdowns. The effect size of lockdowns was not only positive and significant but was also increasing till the end of December 2020.

## 2 Methods

### 2.1 Data

We used three sets of data to conduct our study: mental health claims data including emergency department (ED) claims, COVID-19 cases data, and lockdown dates data.

The mental health data is a large de-identified medical claims corpus provided by Change Healthcare for years of 2019 and 2020. Change Healthcare serves 1 million providers covering 5500 hospitals with 220 million patients (which is roughly two-thirds of the US population) and represents over 50% of private insurance claims across the United States. It covers 51 states/territories and a total of 3141 counties (and equivalent jurisdictions like parishes). The data set includes millions of claims per month from the private insurance marketplace, and some Medicare Advantage programs and Medicaid programs using private insurance carriers, excluding Medicare and Medicaid indemnity claims, which is a limitation in the dataset coverage. More details on the selection method of mental disorders are found in Appendix table A.1

For COVID-19 cases, we considered state-level and county-level cases reported in the United States taken from the New York Times database [49] from the first case date in late January 2020 to December 31, 2020 covering 3218 counties in 51 states/territories. Given that reported cases depend on the testing results, thus, the data is limited by the fact that there was a widespread shortage of available tests in different regions at different times. The undercounts of COVID-19 cases used in this study would only weaken the effect we present, and so fixing the data would only strengthen the resultant effect.

For lockdown data, we used the data from the COVIDVis project ^2^ led by the University of California Berkeley to track policy interventions on state and county levels, in which they depended on government pandemic responses to construct the dataset. We considered the dates of two order types, *shelter-in-place* and *K-12 school closure* in state and county levels. Earliest and latest shelter-in-place orders were on March 14 and April 7, 2020 covering 2598 counties in 43 states. The earliest K-12 school closure was on March 10 and the latest was on April 28, 2020 covering 2465 counties in 39 states. The data is comprehensive, in which states and counties that do not appear in the dataset are considered without official imposed lockdown. We focus on the impact of the initial shutdowns to avoid complications related to re-opening and repeated closures. Given that at some regions people tend to voluntarily isolate themselves at home and limit their trips before official lockdown orders [34], therefore, lockdown dates might be limited to reflect the actual social distancing behavior across regions during the pandemic. However, lockdown dates would better reflect the beginning of persistent social distancing behaviors for a larger population group, which is useful to our study, unlike voluntarily behaviors.

### 2.2 Difference-in-Differences Analysis

To estimate the effects of COVID-19 mitigation policies on mental health patients in county and state levels, we conducted difference-in-differences (DID) analysis, which allows for inferring causality based on parallel trends assumption. For DID analysis we considered mental health patients from the date of September 1, 2019 till December 31, 2020. Our approach leveraged the variation of policy mandated dates in different counties or states with 8 states that did not declare an official lockdown. Accordingly, we constructed both treated and control groups to implement the analysis. We estimated the following regression as our main equation for counties Eq. 1 (same as for states Eq. 2):

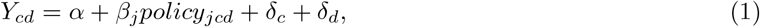

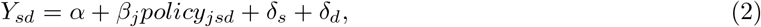

where *Y*_*cd*_ is the outcome (e.g. total mental health patients) in a given county *c* or state *s* on date *d, policy*_*jcd*_ indicates whether policy *j* has been mandated for a county *c* on date *d, β*_*j*_ is the DID interaction coefficient, representing the effect of introducing policy *j*, and *δ*_*c*_ and *δ*_*d*_ are fixed effects for county and date respectively. County fixed effect is included to adjust for time-invariant (independent of time) unobserved county characteristics that might affect the outcome. For example, each county has its local health care system, social capital index, age profile, and socioeconomic status that the fixed effect controls for. Further, date fixed effect *δ*_*d*_ is included to adjust for factors that vary over time, such as COVID-19 rates or social behavioral change.

#### 2.2.1 Control by the evolution of COVID-19 cases

Even though DID avoids the bias encountered in time-invariant factors, the bias of time-varying confounders may still be present [30]. Therefore, we consider the COVID-19 confirmed cases *x*_*cd*_ as a main confounder factor in counties and states and we control for it. We follow [52] to use a Time-Varying Adjusted (TVA) model, based on the assumption that the confounding variable affect both treated and untreated groups regardless of a policy intervention. We measured the interaction of time and the confounding *x*_*cd*_ covariate at county- (Eq. 3) and state-level (Eq. 4)

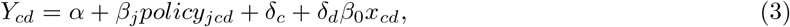

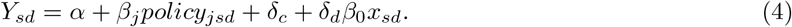

#### 2.2.2 Event-study model

DID models rely on the assumption of parallel pre-treatment trends to exist in both treated and untreated groups. Hence, in the absence of a policy, treated counties or states would evolve similarly as untreated counties or states. To assess equal pre-policy trends, we designed an event-study type model [26]. We calculated *k* periods before policy implementation and used an event-study coefficient to indicate whether an outcome in specific date *d* and county *c* or state *s* is within *k* periods before the policy implementation [44, 19]. We estimated the following regression model for counties (Eq. 5) and states (Eq. 6):

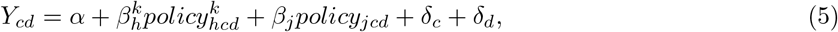

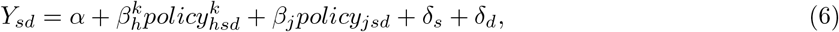

where 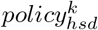, a dummy variable, equals 1 if policy *h* took place *k* periods before the mandate, and zero otherwise. Period *k* is calculated in months, *k* = *{−*6, *−* 5, *−* 4, *−* 2, *−* 1, 0*}* months, and the month of the policy implementation (*k* = 0) is considered as the omitted category. Here, 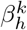 is the event-study coefficient and we included all control variables as defined in equations 1 and 2.

## 3 Results

### 3.1 Descriptive Analysis

Before we delve into the causal DID inference, we report some statistics to describe the data of mental health patients. Among 16.7 million mental health patients in the U.S, the mean age was 38.7 years and 56% were female. As seen in Figure 1, the distribution of mental health patients in states and counties had shifted between 2019 and 2020. The total increase is 22% of all mental health patients of any mental health disorder as seen in Table B.2 in the appendix.

**Figure 1:**
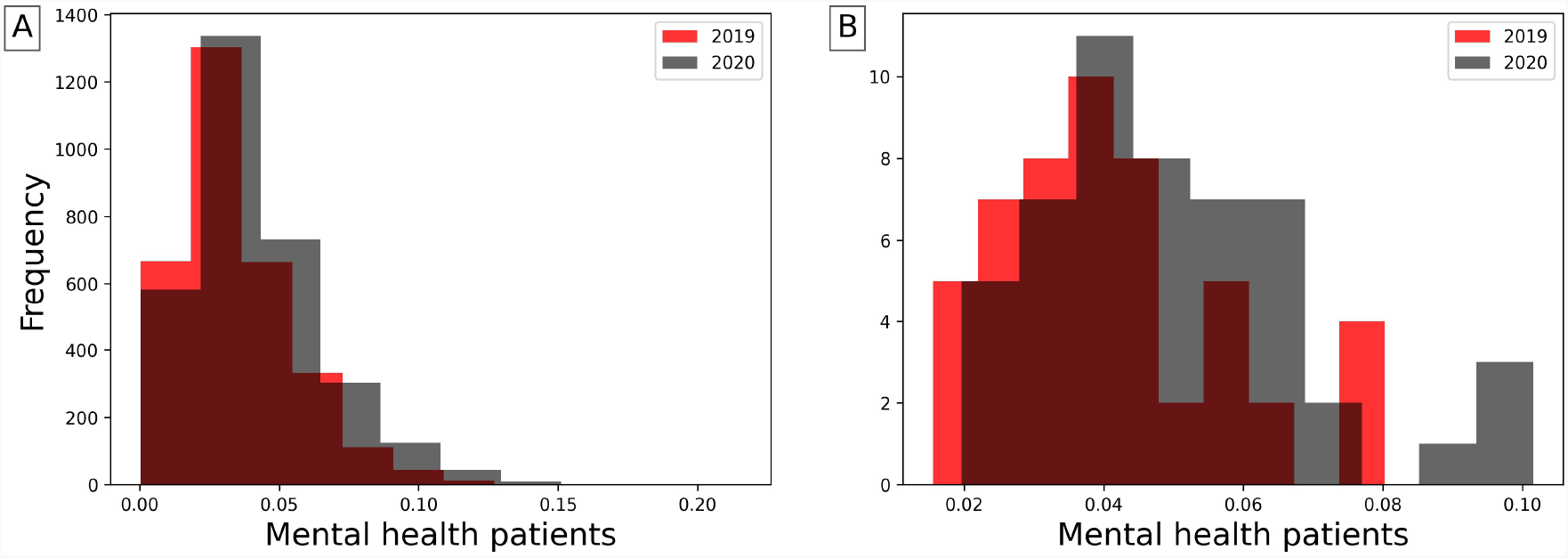
Distributions of mental health patients weighted by regions’ populations in years of 2019 and 2020 in counties (A) and states (B). The total population increase is 22% in 2020

Figure G.6 (in appendix) shows the increasing trend of number of mental health patients, though it decreased between March and April 2020, during lockdown mandates. This supports the findings of [50] where visits of anxiety patients significantly decreased right after strict lockdown took place. An obvious increase was during June 2020, which can be attributed to telemedicine options or relaxed lockdown measures.

#### Parallel Trend Assumption

To apply DID, first we must validate the pre-policy parallel trends assumption. We tested the equality of pre-policy trends for counties and states using Eq. 5 and Eq. 6 respectively. We plot the event-study coefficients for 6 months before policy implementation from the models of stay-at-home and school-closure orders and the corresponding 95 % confidence intervals. Figure D.5 (in appendix) shows that the event-study coefficients are generally non-significant, therefore we cannot reject the null hypothesis of parallel trends. Accordingly, the key assumption of parallel trends of DID is satisfied for both counties and states.

### 3.2 Effects on the Usage of Mental Health Resources

We consider mental health patients for the causal DID inference model from September 1, 2019 to December 31, 2020 to observe the prolonged effects since mental health disorders may appear some time after a trauma [8]. In Tables B.7 (in appendix) and B.8 (in appendix) we summarize the estimated effects based on Eq. 1 and Eq. 2 of stay-at-home and school closure lockdowns on the mental health in counties and states respectively for different population groups with the adjusted results after controlling for COVID-19 cases. Along with regression estimates, we include significance measures of p-value, 95% confidence intervals of standard errors, and R-squared (*R*^2^).

We selected COVID-19 confirmed cases to be a confounding effect and adjusted the regression models using TVA model of Eq. 3 and Eq. 4. Figure 2 confirms COVID-19 effect on mental health patients with significant correlation between both populations (R^2^=0.77, p-value *<* 2*×*10^*−*16^).

**Figure 2:**
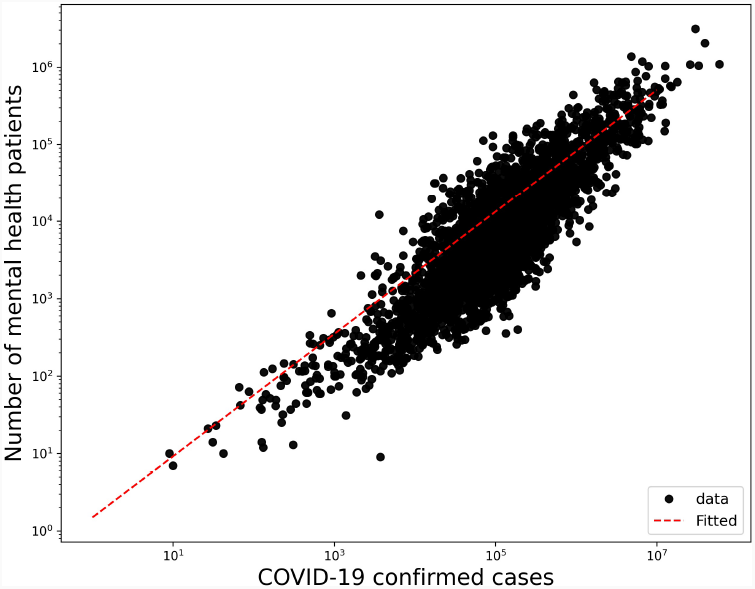
Correlation of mental health patients and COVID-19 confirmed cases in a log-log plot with an increase of 0.043 mental health patients for each confirmed COVID-19 case in counties (R^2^=0.77, p-value *<* 2 *×* 10^*−*16^)

Tables F.13 and F.14 in appendix summarize the estimated effects of Eq. 1 and Eq. 2 at different periods of time *k* where *k*= *{*1,5,9*}* -months after lockdowns, to show the dynamic effect of stay-at-home and school closures in counties and states respectively.

Figures 3 and G.7 (in appendix) show the average differences of mental health patients between counties with lockdown and without lockdown, for stay-at-home and school closure orders respectively. Similarly, Figures G.8 and G.9 in the appendix show the average differences at state-level.

**Figure 3:**
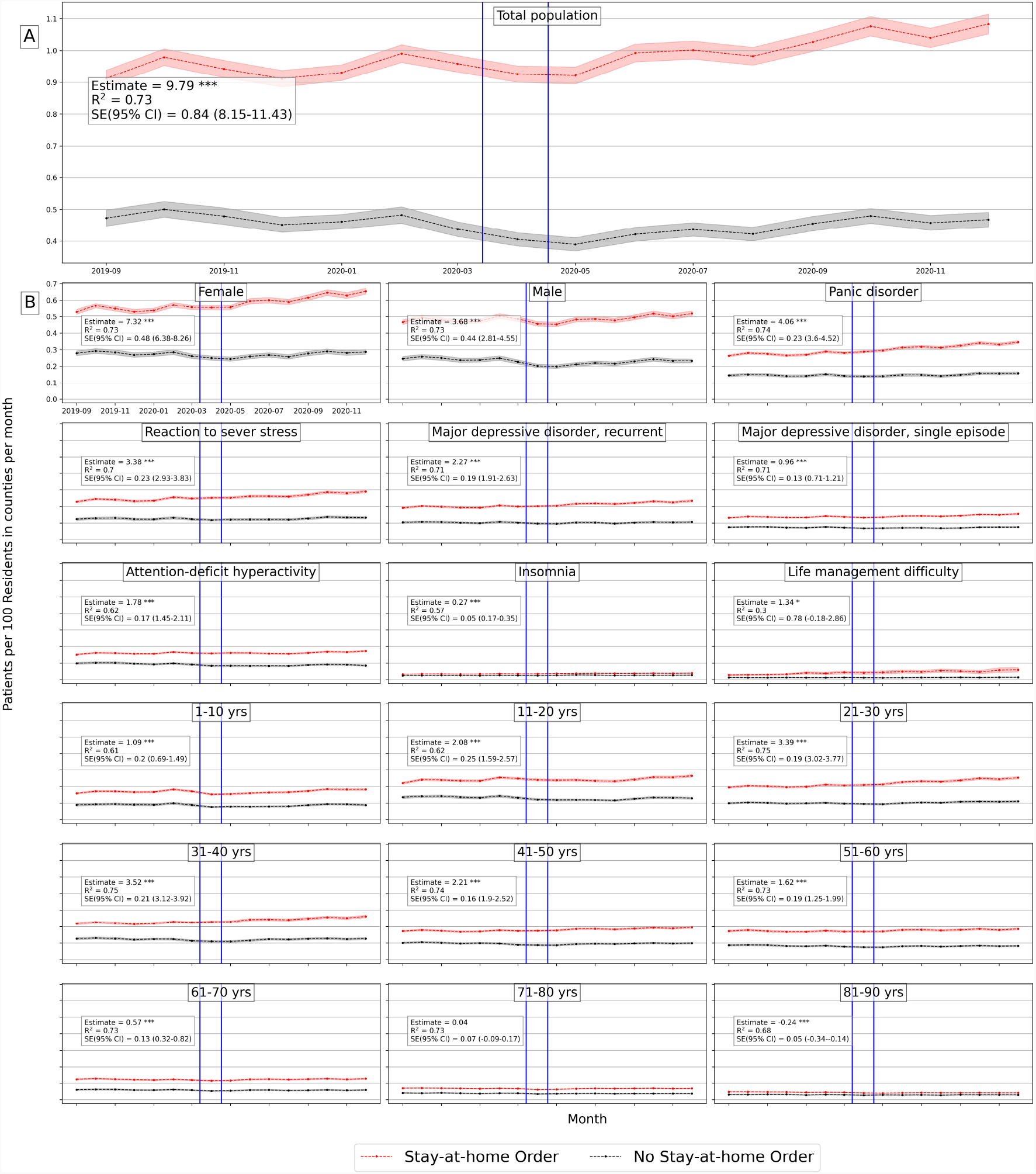
Average number of mental health patients over time (September 2019 - December 2020) in counties with stay-at-home orders and without. Vertical lines show the first stay-at-home order on 3/14/2020 and last 0n 4/07/2020 across United States. Difference-in-differences estimates are included for each population. (Detailed average percentage changes are listed in Table B.3) *\* * * p <* 0.01 *\* * p <* 0.05 *\* p <* 0.1

We will further discuss results for each population group in both counties and states.

#### 3.2.1 Effects on Total Population

Based on Table B.7 (in appendix) there is a significant positive effect of stay-at-home order across counties on the total number of mental health patients, with a mean difference of 9.79 patients between counties with stay-at-home orders and counties without. On average, mental health patients increased by 18.7% but declined by 1% in counties without lockdown (Figure 3). Adjusting for COVID-19 confounding effect preserves the positive effect significant on mental health patient population with a lower effect size of 3.81 estimated mean difference (Table B.7 in appendix). School closure has also a significant, but lower effect on mental health patient population (estimated mean difference = 2.35), with percentage increase of 17% and 16% in counties with closed schools and without respectively (Figure G.7 in appendix), with no significant effect while adjusted for COVID-19 cases.

Similar results are found at the state-level, Table B.8 (in appendix) shows that the effect of stay-at-home order is positively significant for total mental health patients (difference estimate is 466.62 and 439.2 when adjusted) with 22% increase by December 2020 as compared to less than 2% increase in states without lockdown (Figure G.8 in appendix). However, school closures have no significant effect at the state-level.

Table F.13 (in appendix) shows that the lockdown effect size keeps increasing from the first month after the lockdown date until the end of year 2020, for both stay-at-home orders and school closures in counties (Table F.13 in appendix) and states (Table F.14 in appendix).

#### 3.2.2 Gender Effects

In counties, the estimated effect of stay-at-home orders on both females and males are 7.32 (3.32 when adjusted) and 3.68 (1.3 when adjusted) respectively (Table B.7 in appendix). Female patients increased by 24% in counties with stay-at-home orders in comparison with 3% in counties without (Figure 3 in appendix). Male patients declined by 5% in counties without stay-at-home orders. The estimated effect of school closures is significant for females (mean difference = 2.21, and 0.77 when adjusted), but not significant for males (Table B.7 in appendix).

Similarly in states, the estimated mean difference for females is 322.9 (292.37 when adjusted) and for males is 144.1 (187.27 when adjusted) (Table B.8 in appendix). Female patients increased by 29% and 6% in states with stay-at-home orders and without respectively, while male patient numbers decreased in states without stay-at-home lockdown (Figure G.8 in appendix). Similarly, school closures caused significant effect on female patients (estimation = 113.61 and not significant when adjusted) but the effect is not significant on male patients (Table B.8 in appendix).

The effect of both lockdowns on female and male patients kept increasing significantly throughout the year of 2020 in counties and states (Tables F.13, F.14 in appendix)

#### 3.2.3 Diagnosis Effects

We selected top five mental disorders (e.g. *panic disorder*) that peaked in 2020, and other disorders of interest (*insomnia* and *life management difficulty*) to investigate the effect of lockdowns on patient populations for specific diagnosis.

In counties, all disorders were positively and significantly affected by stay-at-home orders and mostly by school closures with lower effect sizes. Patients diagnosed with *panic disorder* (ICD-10: F41) had the largest difference effect among other mental illnesses and increased in both county groups (31.8% vs 8.88%) with estimated effect of 4.06 (1.51 when adjusted). Patients with *attention-deficit hyperactivity disorder* (ICD-10: F90) decreased in counties without stay-at-home order by -13.6% with estimated effect of 1.78 (1.55 when adjusted). Patients diagnosed with *life management difficulty* (ICD-10: Z73) (i.e. burnout) doubled in counties with stay-at-home orders, compared to 10% increase in counties without such lockdown with estimated effect 1.34 (1.55 when adjusted) (Figure 3). Unlikely, patients with *insomnia*, with significant estimated effect of -0.09 when adjusted, increased more in counties without school closures by 24% compared to 17% in counties with closures. Patients diagnosed with *life management difficulty* disorder increased more in counties without school closures by 127.85% compared with 94.64% with closures, and the estimated effect is -5.14 and not significant when adjusted (Table B.7 and Figure G.7 in appendix).

Similarly at the state-level, *panic disorder* (ICD-10: F41) increased by 38.4% in states with stay-at-home orders (Figure G.8 in appendix) and had the largest difference effect size with mean difference of 170.8 and 136.75 when adjusted (Table B.8 in appendix). Patients with *life management difficulty* increased more in states without a school closure by 161.49% compared to 123.36% in states with closures with estimated effects of -14.35 and -14.88 when adjusted.

Over time, the effect of stay-at-home order kept increasing significantly for all selected mental disorders across counties (Table F.13 in appendix) and states (Table F.14 in appendix). While school closure effect is significantly increasing for most diagnosis except for *life management difficulty* diagnosis where the effect kept declining.

#### 3.2.4 Age Effects

At the county-level, for most age groups, both lockdowns have positive significant effects on mental health patient population, except the eldest age group (*>* 80 yrs old), which showed a negative effect. Based on Table B.7 (in appendix), the two largest significant differences were for adults between 31 and 40 years old and adults between 21 and 30 years old. Adults in their thirties increased by 20.47% in counties with stay-at-home order but declined by -0.1% in counties without, with mean difference of 3.52 (1.74 when adjusted). Adults in their twenties increased more in counties with stay-at-home orders by 30.01% compared to 11% in counties without, with estimated effect of 3.39 (1.3 when adjusted). Young patients under 11 and adolescent patients under 21 decreased in counties without stay-at-home orders by -1.2% and -4.5% respectively. The estimated effect for children under 11 is 1.09, but not significant when adjusted, and for adolescents (11 to 20) is 2.08 and 0.7 when adjusted (Figure 3).

Similarly, school closures affected patients in their thirties but with lower mean differences of 0.88 (not significant when adjusted) (Table B.8 in appendix). They increased by 18.75 vs. 18.62 in both regions with and without closures respectively. While Teenager and adolescent (11 to 20) patients increased more in counties with school closures by 27.16%, compared to 19.17% in counties without closures, with estimated effect 0.6 (not significant when adjusted) (Figure G.7 in appendix).

Similar observations are found at the state-level based on Table B.8 (in appendix). For most age groups both stay-at-home and school closure orders show significant positive effect, with larger effect sizes for people aged less than 40 years old. Mental health patients who are in their thirties increased by 28% and 1% in states with stay-at-home order and without respectively. Similarly, patients in their twenties increased by 40% and 15% in states with stay-at-home order and without respectively (Figure G.8 in appendix).

The effect size of both lockdowns on most age groups kept increasing significantly throughout the year of 2020. Children less than 11 years old had the largest change of estimation size, which indicates a greater effect on children appeared later on in counties with stay-at-home orders (Table F.13 in appendix).

### 3.3 Effects on Urgent Treatment-Seeking

Emergency department (ED) visits have been considered to reflect the emergent need to seek a mental health facility during COVID-19 pandemic. The effects on ED visits have shown similar trend as previous results. The effect of stay-at-home order on total population is positive and significant with magnitude of 0.29 weighted by population on state-level (See Figure 4, and Figure G.10 for the rest of the groups in appendix). Similarly, the effect of school closure is positive and significant with value of 0.12 weighted by state population (See Figure G.11 in appendix). Most of the other groups also showed positive and significant effects on both county and state levels. Table C.10 (in appendix) shows the effects of both orders on all the selected groups on state-level, and Table C.9 (in appendix) on county-level.

**Figure 4:**
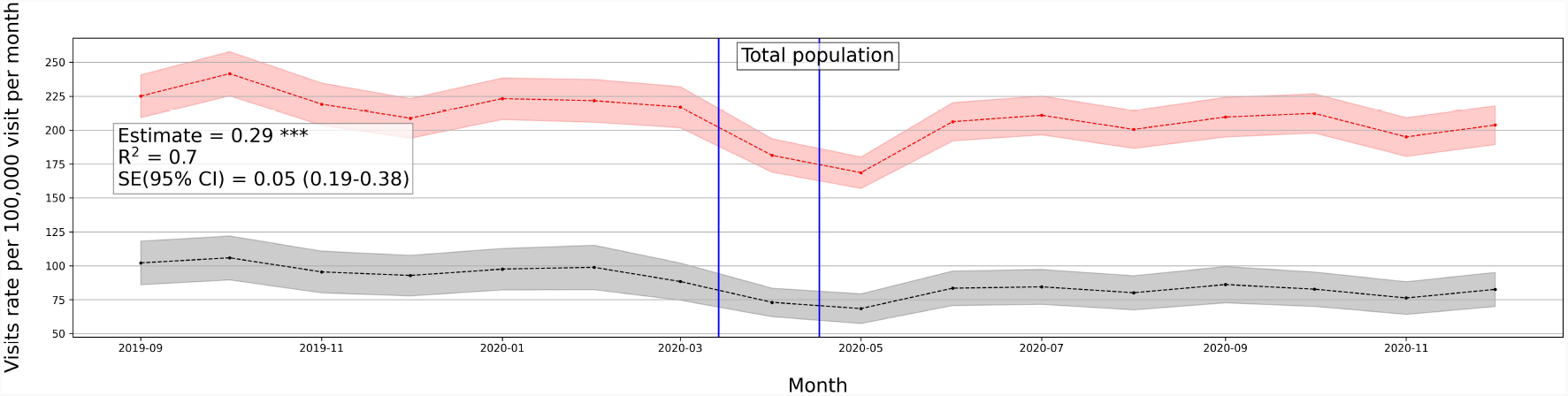
Average number of mental health ED visits over time (September 2019 - December 2020) in counties with stay-at-home orders and without. Vertical lines show the first stay-at-home order on 3/14/2020 and last on 4/07/2020 across United States. Difference-in-differences estimates are included for each population. *\* * * p <* 0.01 *\* * p <* 0.05 *\* p <* 0.1

### 3.4 Robustness Check

Given the differences in regions with respect to the number of hospitals, facilities and patients, we conducted robustness checks of our main analysis to show that dropping multiple states does not change the estimates, and that our results are not driven by specific regions. We dropped New York and Ohio states which both were the most two states with large patients volume relative to population. The estimates remained robust, significant and positive (Table E.11 in appendix). We also added all 2019 samples to expand the control group and the pre-intervention period. The causality inferred from our analysis stayed significant and positive with this expansion (Table E.12 in appendix).

## 4 Discussion

Early in March 2020, non-pharmaceutical interventions, such as social distancing policies, were imposed around the world to contain the spread of COVID-19 and proved to reduce the number of COVID-19 cases and fatalities [23, 24, 12]. Mitigation policies come with both costs and benefits, which may be further analyzed to help determine the optimal time to release or stop a policy intervention [33]. Prior research showed significant mental health degradation associated with COVID-19 pandemic [17, 27, 2, 7, 19], however no research investigated the causal relation between COVID-19 mitigation policies and the usage of mental health resources. Considering the effects on the usage of mental health resources can further reflect the economic and health costs brought by the pandemic interventions. In our study, using large-scale medical claims data, we estimated the effects of lockdowns on the usage of mental health facilities and the prevalence of mental health issues at the state-and county-levels in the United States.

Our findings demonstrate a statistically significant causal effect of lockdown measures (stay-at-home and school closure orders) on the usage of mental health facilities represented by increasing number of issued medical claims for mental health appointments during COVID-19 pandemic. Also, ED visits were statistically significant and positive in locked-down regions which reflect the increase of emergent mental help-seeking due to the COVID-19 lockdowns. Results further emphasize the cost brought by extra months of lockdowns, in which effect sizes keep increasing through the end of 2020 in both mental health visits and ED visits.

Some sub-population groups were exposed to larger deterioration effect than other groups, such as female and adolescent groups.

Given the various intertwined events and causes during the COVID-19 pandemic, our analysis is limited by several factors. First, it is important to point out that the adoption of lockdowns across states did not happen at random. Differences of shutdown orders’ timings and adoption across regions were associated with the differences in COVID-19 confirmed cases and fatality rates across those regions [29, 35] and the differences in their health systems capacity [28]. Also, there exist other political, economical, and institutional factors that affect the adoption of COVID-19 measures and their strictness level across countries [20]. Even though the lockdown timing may be affected by regional factors related to the virus, such as number of cases or institutional factors, however, there is no reason to believe that lockdown timing was affected the prevalence of mental health in regions. Given that, we have also encountered regional fixed effect in our model to adjust for regional differences. Second, though mental illnesses have a negative economic impact [43], the opposite is true as well, in which economic disadvantage may lead to a greater mental illness [32]. During COVID-19, there have been negative consequences on individuals in different industry sectors who were more likely to lose their jobs due to the lockdown measures [18] with significant employment loss in occupations that require interpersonal contact [39]. Therefore, the loss of employment due to shutdowns may have a confounding effect on increased mental health issues.

In addition, the medical claims used in this study do not cover Medicare and Medicaid health insurance programs which creates a limitation on our data. Medicare covers most aged and disabled population across the US, while Medicare covers a wider range of population including low-income beneficiaries covering 30% of US population [31]. This limitation would impact the representativeness of results, since our data misses some population groups in the US.

Despite the mentioned limitations, our results provide important policy implications from economic and social impacts. There is a notable mental health cost brought by non-pharmaceutical interventions, especially interventions that are extended to longer duration. Our results suggest that there should be considerations to the mental health cost through ensuring mental health treatment capacity.

Furthermore, we showed that number of patients had dropped right after lockdowns and then progressively increased in June and July 2020, supporting the findings of [50, 41]. This suggests that people with mental health afflictions did not have the ability to seek immediate care during restrictive lockdowns. Findings suggest that policy interventions should be accompanied with strategies that facilitate mental health treatment despite restrictive lockdowns, in order to avoid the exacerbated effect of delayed treatment.

## Data Availability

1. Medical claims data is confidential, provided by Change Healthcare, LLC
2. NYTimes COVID-19 data (publicly available)
3. COVID-19 government responses data (publicly available)

https://github.com/nytimes/covid-19-data

https://github.com/covidvis/covid19-vis/tree/master/data

## 5 Acknowledgment

We thank the Change Healthcare team, Craig Midgett, Mina Atia, Andrew Harris, Anil Konda, Tim Suther, and Jaideep Kulkarni for facilitating our access to medical claims data and for their help in large-scale analysis.

## Appendices

**Appendix A Table for Syndrome Definitions and Diagnosis Codes**

**Appendix B Effects of COVID-19 Lockdowns on Mental Health Resource Usage**

**Appendix C Effects of COVID-19 Lockdowns on Mental Health ED Visits**

**Appendix D Event-Study Pre-trend Check**

**Appendix E Robustness Check**

**Appendix F Extended Effects of COVID-19 Lockdowns**

**Appendix G Figures**

**Table A.1:**
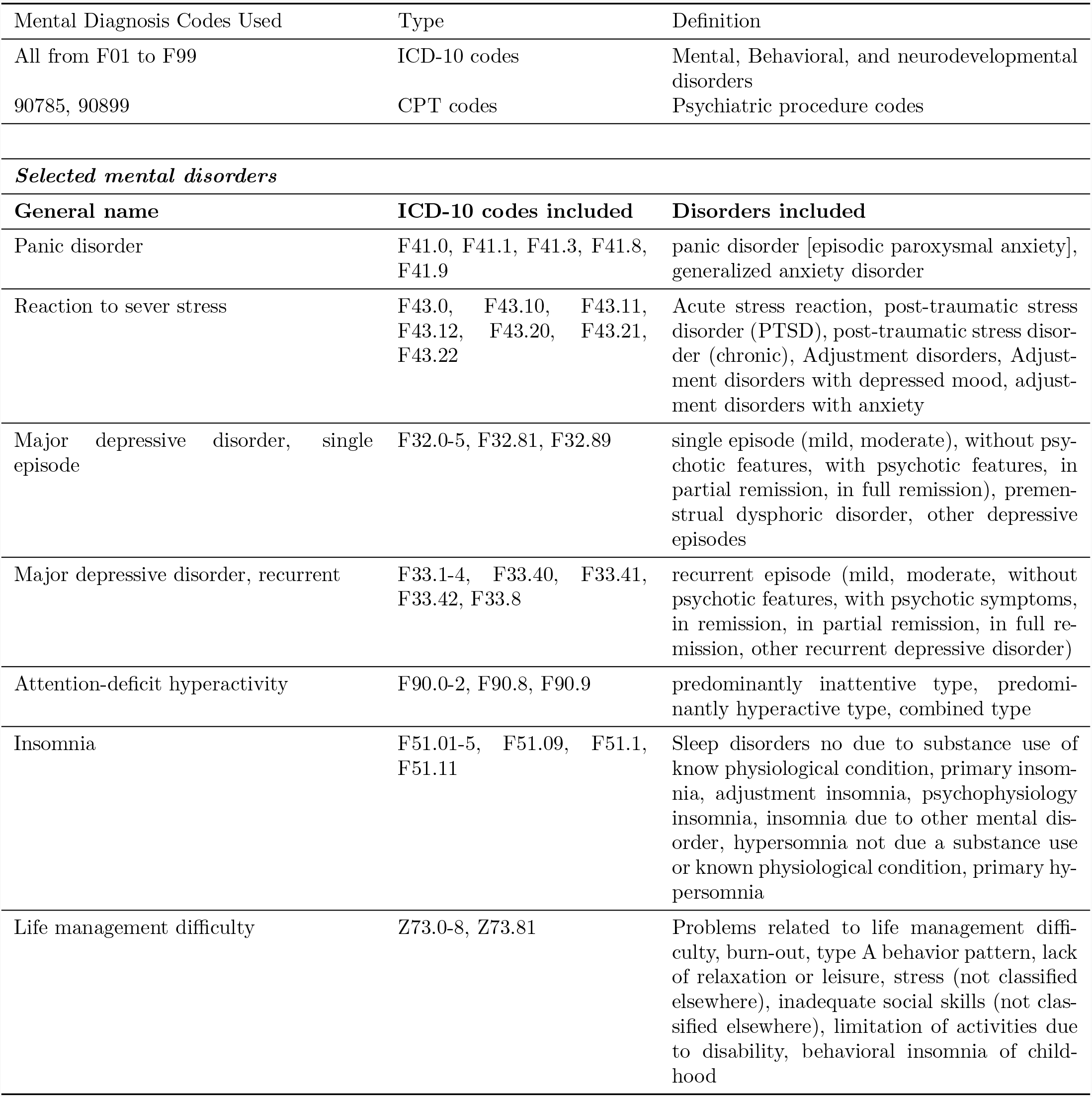
Definitions and descriptions of the mental health codes used to select medical claims of interest

| Mental Diagnosis Codes Used | Type | Definition |
| --- | --- | --- |
| All from F01 to F99 | ICD-10 codes | Mental, Behavioral, and neurodevelopmental disorders |
| 90785, 90899 | CPT codes | Psychiatric procedure codes |
| ***Selected mental disorders*** |  |  |
| General name | ICD-10 codes included | Disorders included |
| Panic disorder | F41.0, F41.1, F41.3, F41.8, F41.9 | panic disorder [episodic paroxysmal anxiety], generalized anxiety disorder |
| Reaction to severe stress | F43.0, F43.10, F43.11, F43.12, F43.20, F43.21, F43.22 | Acute stress reaction, post-traumatic stress disorder (PTSD), post-traumatic stress disorder (chronic), Adjustment disorders, Adjustment disorders with depressed mood, adjustment disorders with anxiety |
| Major depressive disorder, single episode | F32.0-5, F32.81, F32.89 | single episode (mild, moderate), without psychotic features, with psychotic features, in partial remission, in full remission), premenstrual dysphoric disorder, other depressive episodes |
| Major depressive disorder, recurrent | F33.1-4, F33.40, F33.41, F33.42, F33.8 | recurrent episode (mild, moderate, without psychotic features, with psychotic symptoms, in remission, in partial remission, in full remission, other recurrent depressive disorder) |
| Attention-deficit hyperactivity | F90.0-2, F90.8, F90.9 | predominantly inattentive type, predominantly hyperactive type, combined type |
| Insomnia | F51.01-5, F51.09, F51.1, F51.11 | Sleep disorders not due to substance use or known physiological condition, primary insomnia, adjustment insomnia, psychophysiology insomnia, insomnia due to other mental disorder, hypersomnia not due to substance use or known physiological condition, primary hypersomnia |
| Life management difficulty | Z73.0-8, Z73.81 | Problems related to life management difficulty, burn-out, type A behavior pattern, lack of relaxation or leisure, stress (not classified elsewhere), inadequate social skills (not classified elsewhere), limitation of activities due to disability, behavioral insomnia of childhood |

**Table B.2:** Percentage change between 2019 and 2020 mental health patients among population groups

| Group | Change |
| --- | --- |
| Total Population | 22% |
| Female | 20% |
| Male | 14% |
| Panic disorder | 33% |
| Reaction to sever stress | 31% |
| Major depressive disorder, recurrent | 25% |
| Major depressive disorder, single episode | 24% |
| Attention-deficit hyperactivity | 19% |
| Insomnia | 45% |
| Life management difficulty | 73% |
| 1-10 yrs | 28% |
| 11-20 yrs | 20% |
| 21-30 yrs | 28% |
| 31-40 yrs | 24% |
| 41-50 yrs | 20% |
| 51-60 yrs | 17% |
| 61-70 yrs | 15% |
| 71-80 yrs | 16% |
| 81-90 yrs | 6% |

**Table B.3:** (Supplement table for Figures 3 and G.7). Percentage change of monthly average mental health population in counties with and without lockdowns between early September 2019 to end of December 2020

| Group | Stay-at-home |  | School Closure |  |
| --- | --- | --- | --- | --- |
|  | No lockdown | Lockdown | No lockdown | Lockdown |
| Total Population | -1.01% | 18.73% | 16.16% | 17.09% |
| Female | 2.88% | 24.13% | 20.89% | 22.53% |
| Male | -5.33% | 11.05% | 11.88% | 9.06% |
| Panic disorder | 8.88% | 31.18% | 28.49% | 29.17% |
| Reaction to sever stress | 7.35% | 26.86% | 25.82% | 24.83% |
| Major depressive disorder, recurrent | 1.95% | 22.29% | 23.97% | 19.63% |
| Major depressive disorder, single episode | 1.02% | 18.00% | 17.10% | 16.10% |
| Attention-deficit hyperactivity | -13.66% | 14.04% | 10.83% | 11.28% |
| Insomnia | 10.99% | 19.29% | 24.08% | 17.49% |
| Life management difficulty | 10.24% | 111.76% | 127.85% | 94.64% |
| 1-10 yrs | -1.17% | 14.84% | 13.19% | 13.09% |
| 11-20 yrs | -4.54% | 19.85% | 14.70% | 17.63% |
| 21-30 yrs | 10.93% | 30.01% | 30.79% | 27.95% |
| 31-40 yrs | -0.10% | 20.47% | 18.62% | 18.75% |
| 41-50 yrs | -1.42% | 13.57% | 11.91% | 12.55% |
| 51-60 yrs | -5.16% | 7.68% | 10.69% | 6.13% |
| 61-70 yrs | -2.95% | 2.89% | 9.37% | 1.60% |
| 71-80 yrs | -8.34% | -2.75% | -0.29% | -3.68% |
| 81-90 yrs | -6.41% | -12.00% | -5.02% | -12.88% |

**Table B.4:** (Supplement table for Figures G.8 and G.9). Percentage change of monthly average mental health population in states with and without lockdowns between early September 2019 to end of December 2020

| Group | Stay-at-home |  | School Closure |  |
| --- | --- | --- | --- | --- |
|  | No lockdown | Lockdown | No lockdown | Lockdown |
| Total Population | 1.67% | 21.87% | 18.40% | 20.45% |
| Female | 5.91% | 28.86% | 24.00% | 27.42% |
| Male | -3.42% | 13.93% | 13.10% | 12.24% |
| Panic disorder | 15.80% | 38.38% | 35.25% | 36.56% |
| Reaction to sever stress | 15.64% | 36.83% | 34.08% | 35.07% |
| Major depressive disorder, recurrent | 9.38% | 30.51% | 31.34% | 28.07% |
| Major depressive disorder, single episode | 6.29% | 22.95% | 19.62% | 21.89% |
| Attention-deficit hyperactivity | -9.85% | 21.57% | 13.74% | 19.47% |
| Insomnia | 23.07% | 37.47% | 39.43% | 35.55% |
| Life management difficulty | 12.20% | 145.77% | 161.49% | 123.36% |
| 1-10 yrs | 6.21% | 26.91% | 18.26% | 26.29% |
| 11-20 yrs | 4.11% | 28.00% | 19.17% | 27.16% |
| 21-30 yrs | 15.48% | 39.28% | 36.46% | 37.37% |
| 31-40 yrs | 1.36% | 27.60% | 21.55% | 25.93% |
| 41-50 yrs | -1.55% | 18.79% | 14.77% | 17.35% |
| 51-60 yrs | -7.41% | 11.48% | 10.75% | 9.75% |
| 61-70 yrs | -10.83% | 3.75% | 4.49% | 2.39% |
| 71-80 yrs | -10.55% | -3.66% | -2.05% | -4.66% |
| 81-90 yrs | -15.99% | -18.71% | -16.15% | -19.00% |

**Table B.5:**
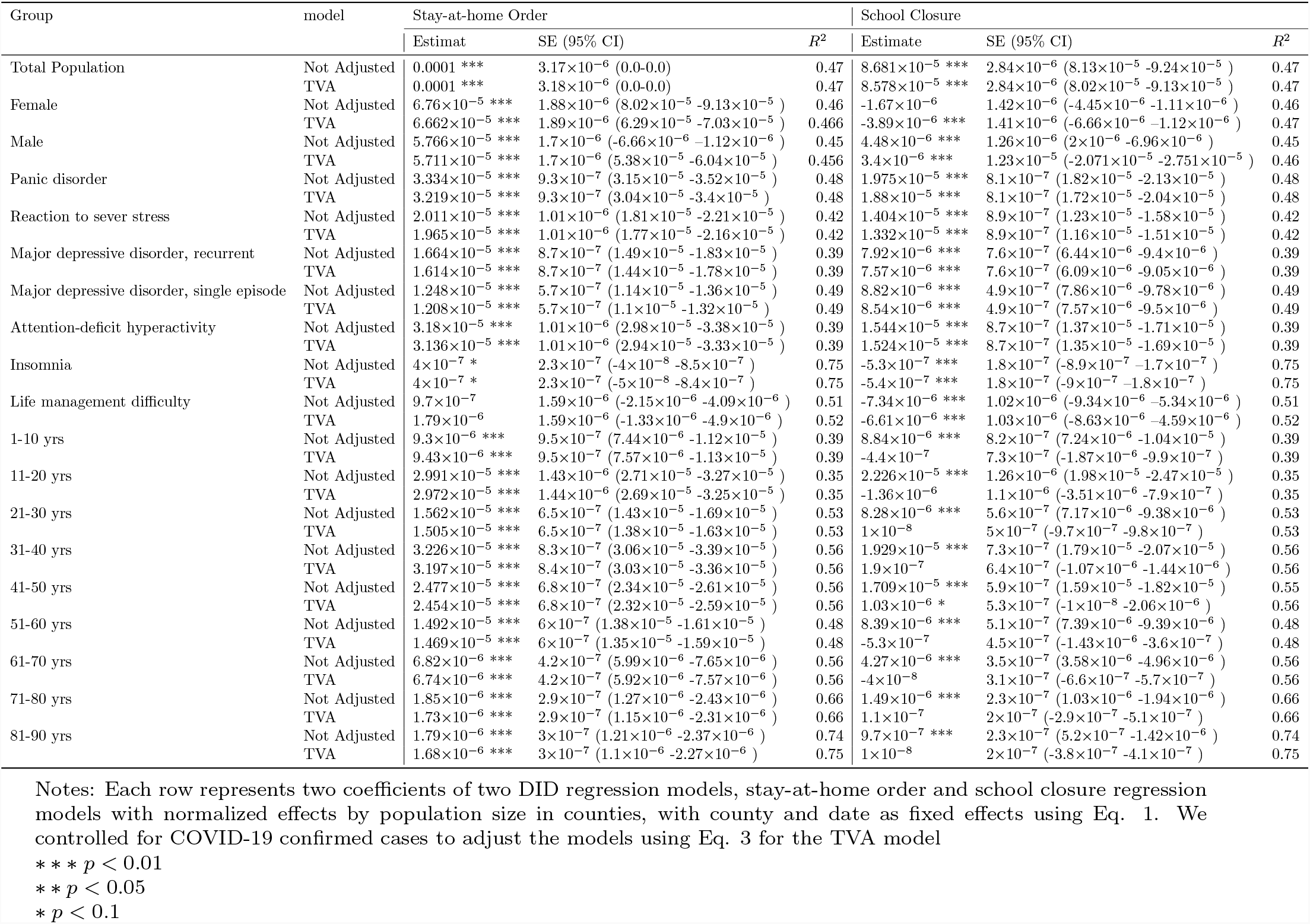
Effects of lockdown interventions on mental health of different population groups in counties weighted by county-level population size

**Table B.6:**
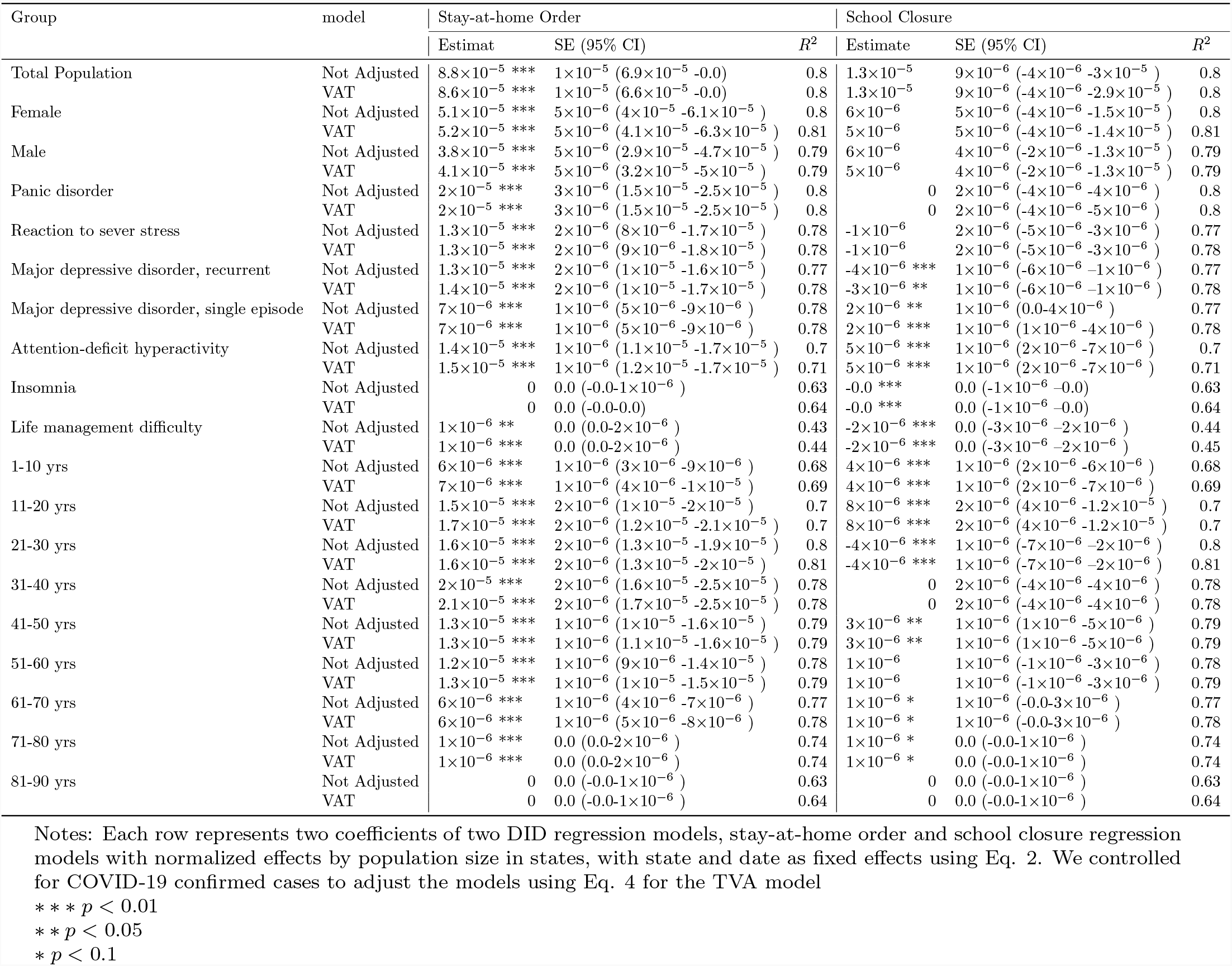
Effects of lockdown interventions on mental health of different population groups in states weighted by state-level population size

**Table B.7:** Effects of lockdown interventions on mental health of different population groups in counties

| Group | model | Stay-at-home Order |  |  | School Closure |  |  |
| --- | --- | --- | --- | --- | --- | --- | --- |
|  |  | Estimat | SE (95% CI) | R<sup>2</sup> | Estimate | SE (95% CI) | R<sup>2</sup> |
| Total Population | Not Adjusted | 9.79 *** | 0.84 (8.15-11.43) | 0.73 | 2.35 *** | 0.74 (0.89-3.81) | 0.73 |
|  | TVA | 3.81 *** | 0.76 (2.33-5.3) | 0.78 | -0.11 | 0.68 (-1.44-1.21) | 0.78 |
| Female | Not Adjusted | 7.32 *** | 0.48 (6.38-8.26) | 0.73 | 2.21 *** | 0.42 (1.38-3.03) | 0.73 |
|  | TVA | 3.32 *** | 0.43 (2.47-4.17) | 0.78 | 0.77 * | 0.4 (-0.0-1.55) | 0.77 |
| Male | Not Adjusted | 3.68 *** | 0.44 (2.81-4.55) | 0.73 | 0.04 | 0.39 (-0.72-0.81) | 0.73 |
|  | TVA | 1.35 *** | 0.41 (0.55-2.15) | 0.77 | 0.22 | 0.3 (-0.37-0.82) | 0.77 |
| Panic disorder | Not Adjusted | 4.06 *** | 0.23 (3.6-4.52) | 0.74 | 1.25 *** | 0.2 (0.84-1.65) | 0.74 |
|  | TVA | 1.51 *** | 0.21(1.1-1.92) | 0.8 | 0.13 | 0.19(-0.24-0.5) | 0.78 |
| Reaction to sever stress | Not Adjusted | 3.38 *** | 0.23 (2.93-3.83) | 0.7 | 0.65 *** | 0.2 (0.26-1.05) | 0.7 |
|  | TVA | 1.33 *** | 0.21(0.92-1.75) | 0.74 | -0.15 | 0.19(-0.53-0.23) | 0.73 |
| Major depressive disorder, recurrent | Not Adjusted | 2.27 *** | 0.19 (1.91-2.63) | 0.71 | 0.36 ** | 0.16 (0.04-0.68) | 0.71 |
|  | TVA | 0.73 *** | 0.17(0.39-1.06) | 0.76 | -0.35 ** | 0.15(-0.65-0.05) | 0.75 |
| Major depressive disorder, single episode | Not Adjusted | 0.96 *** | 0.13 (0.71-1.21) | 0.71 | 0.09 | 0.11 (-0.12-0.31) | 0.71 |
|  | TVA | 0.16 | 0.12(-0.07-0.39) | 0.75 | -0.2 * | 0.11(-0.41-0.01) | 0.73 |
| Attention-deficit hyperactivity | Not Adjusted | 1.78 *** | 0.17 (1.45-2.11) | 0.62 | 0.36 ** | 0.14 (0.08-0.64) | 0.62 |
|  | TVA | 0.95 *** | 0.16(0.64-1.27) | 0.65 | 0.05 | 0.14(-0.22-0.32) | 0.65 |
| Insomnia | Not Adjusted | 0.27 *** | 0.05 (0.17-0.35) | 0.57 | 0.03 | 0.04 (-0.05-0.1) | 0.57 |
|  | TVA | 0.11 ** | 0.04(0.02-0.2) | 0.59 | -0.09 ** | 0.04(-0.16-0.01) | 0.59 |
| Life management difficulty | Not Adjusted | 1.34 * | 0.78 (-0.18-2.86) | 0.3 | -5.14 *** | 0.5 (-6.12-4.17) | 0.3 |
|  | TVA | 1.55 ** | 0.78(0.02-3.09) | 0.3 | -5.09 *** | 0.5(-6.08-4.11) | 0.3 |
| 1-10 yrs | Not Adjusted | 1.09 *** | 0.2 (0.69-1.49) | 0.61 | 0.17 | 0.18 (-0.17-0.51) | 0.61 |
|  | TVA | 0.07 | 0.19(-0.31-0.45) | 0.66 | -0.48 *** | 0.15(-0.77-0.19) | 0.66 |
| 11-20 yrs | Not Adjusted | 2.08 *** | 0.25 (1.59-2.57) | 0.62 | 0.6 *** | 0.22 (0.17-1.02) | 0.62 |
|  | TVA | 0.7 *** | 0.24(0.24-1.16) | 0.66 | -0.17 | 0.18(-0.52-0.18) | 0.66 |
| 21-30 yrs | Not Adjusted | 3.39 *** | 0.19 (3.02-3.77) | 0.75 | 0.78 *** | 0.17 (0.46-1.11) | 0.75 |
|  | TVA | 1.3 *** | 0.17(0.96-1.64) | 0.8 | -0.61 *** | 0.13(-0.87-0.35) | 0.8 |
| 31-40 yrs | Not Adjusted | 3.52 *** | 0.21 (3.12-3.92) | 0.75 | 0.88 *** | 0.18 (0.53-1.23) | 0.75 |
|  | TVA | 1.74 *** | 0.19(1.37-2.12) | 0.79 | 0.07 | 0.14(-0.22-0.35) | 0.79 |
| 41-50 yrs | Not Adjusted | 2.21 *** | 0.16 (1.9-2.52) | 0.74 | 0.72 *** | 0.14 (0.45-0.99) | 0.74 |
|  | TVA | 1.13 *** | 0.15(0.83-1.42) | 0.78 | 0.1 | 0.12(-0.12-0.33) | 0.78 |
| 51-60 yrs | Not Adjusted | 1.62 *** | 0.19 (1.25-1.99) | 0.73 | 0.2 | 0.16 (-0.11-0.52) | 0.73 |
|  | TVA | 0.66 *** | 0.18(0.31-1.01) | 0.75 | -0.21 | 0.14(-0.48-0.06) | 0.75 |
| 61-70 yrs | Not Adjusted | 0.57 *** | 0.13 (0.32-0.82) | 0.73 | -0.11 | 0.11 (-0.32-0.1) | 0.73 |
|  | TVA | 0.1 | 0.12(-0.13-0.34) | 0.76 | -0.05 | 0.09(-0.22-0.13) | 0.76 |
| 71-80 yrs | Not Adjusted | 0.04 | 0.07 (-0.09-0.17) | 0.73 | -0.07 | 0.05 (-0.17-0.04) | 0.73 |
|  | TVA | -0.08 | 0.06(-0.21-0.04) | 0.76 | 0.05 | 0.04(-0.04-0.13) | 0.76 |
| 81-90 yrs | Not Adjusted | -0.24 *** | 0.05 (-0.34-0.14) | 0.68 | -0.25 *** | 0.04 (-0.33-0.17) | 0.68 |
|  | TVA | -0.18 *** | 0.05(-0.28-0.09) | 0.71 | -0.15 *** | 0.03(-0.22-0.09) | 0.71 |

Notes: Each row represents two coefficients of two DID regression models, stay-at-home order and school closure regression models with raw total population in counties, with county and date as fixed effects using Eq. 1. We controlled for COVID-19 confirmed cases to adjust the models using Eq. 3 for the TVA model
\*\*\* $p < 0.01$
\*\* $p < 0.05$
\* $p < 0.1$

**Table B.8:** Effects of lockdown interventions on mental health of different population groups in states

| Group | model | Stay-at-home Order |  |  | School Closure |  |  |
| --- | --- | --- | --- | --- | --- | --- | --- |
|  |  | Estimate | SE (95% CI) | R<sup>2</sup> | Estimate | SE (95% CI) | R<sup>2</sup> |
| Total Population | Not Adjusted | 466.62 *** | 102.86 (265.01-668.23) | 0.78 | 117.18 | 90.37 (-59.95-294.32) | 0.78 |
|  | TVA | 439.19 *** | 94.88 (253.22-625.17) | 0.82 | 82.93 | 83.03 (-79.82-245.68) | 0.82 |
| Female | Not Adjusted | 322.89 *** | 55.02 (215.06-430.73) | 0.79 | 113.61 ** | 48.35 (18.85-208.38) | 0.79 |
|  | TVA | 292.37 *** | 51.09 (192.22-392.51) | 0.82 | 89.99 ** | 44.57 (2.63-177.35) | 0.82 |
| Male | Not Adjusted | 144.06 *** | 48.1 (49.78-238.34) | 0.78 | -11.9 | 42.25 (-94.72-70.91) | 0.78 |
|  | TVA | 187.27 *** | 44.83 (99.4-275.14) | 0.82 | 18.36 | 39.1 (-58.28-95.0) | 0.82 |
| Panic disorder | Not Adjusted | 170.79 *** | 23.48 (124.76-216.81) | 0.8 | 71.45 *** | 20.64 (31.0-111.9) | 0.8 |
|  | TVA | 137.75 *** | 21.71 (95.2-180.3) | 0.83 | 47.78 ** | 18.94 (10.65-84.91) | 0.83 |
| Reaction to sever stress | Not Adjusted | 125.04 *** | 18.98 (87.85-162.24) | 0.77 | 36.31 ** | 16.68 (3.62-69.0) | 0.77 |
|  | TVA | 110.54 *** | 17.98 (75.3-145.79) | 0.81 | 25.02 | 15.7 (-5.75-55.78) | 0.81 |
| Major depressive disorder, recurrent | Not Adjusted | 88.15 *** | 15.41 (57.93-118.36) | 0.78 | 24.53 * | 13.54 (-2.02-51.07) | 0.78 |
|  | TVA | 74.41 *** | 14.13 (46.71-102.12) | 0.82 | 15.01 | 12.33 (-9.17-39.19) | 0.82 |
| Major depressive disorder, single episode | Not Adjusted | 35.63 *** | 10.04 (15.95-55.32) | 0.78 | 8.82 | 8.82 (-8.46-26.11) | 0.78 |
|  | TVA | 36.61 *** | 9.46 (18.06-55.15) | 0.81 | 9.91 | 8.25 (-6.27-26.09) | 0.81 |
| Attention-deficit hyperactivity | Not Adjusted | 59.13 *** | 12.48 (34.67-83.59) | 0.74 | 15.16 | 10.94 (-6.27-36.59) | 0.74 |
|  | TVA | 57.42 *** | 11.53 (34.82-80.01) | 0.79 | 16.87 * | 10.04 (-2.81-36.55) | 0.79 |
| Insomnia | Not Adjusted | 5.83 *** | 1.27 (3.34-8.33) | 0.76 | 2.32 ** | 1.09 (0.18-4.47) | 0.76 |
|  | TVA | 0.17 | 1.17 (-2.12-2.46) | 0.81 | -1.51 | 1.0 (-3.47-0.44) | 0.81 |
| Life management difficulty | Not Adjusted | 6.51 ** | 2.55 (1.51-11.51) | 0.42 | -14.35 *** | 1.96 (-18.19-10.51) | 0.43 |
|  | TVA | 6.69 ** | 2.58 (1.62-11.76) | 0.44 | -14.88 *** | 1.99 (-18.78-10.98) | 0.44 |
| 1-10 yrs | Not Adjusted | 29.48 ** | 14.12 (1.79-57.16) | 0.72 | 9 | 12.37 (-15.24-33.25) | 0.72 |
|  | TVA | 27.1 ** | 12.7 (2.21-51.98) | 0.78 | 10.66 | 11.05 (-11.0-32.32) | 0.78 |
| 11-20 yrs | Not Adjusted | 82.17 *** | 21.12 (40.77-123.57) | 0.73 | 27.29 | 18.55 (-9.06-63.65) | 0.73 |
|  | TVA | 83.2 *** | 19.67 (44.64-121.75) | 0.78 | 30.4 * | 17.16 (-3.24-64.03) | 0.78 |
| 21-30 yrs | Not Adjusted | 112.63 *** | 15.57 (82.11-143.14) | 0.79 | 28.11 ** | 13.68 (1.29-54.92) | 0.79 |
|  | TVA | 101.35 *** | 14.81 (72.31-130.38) | 0.82 | 19.2 | 12.93 (-6.14-44.54) | 0.82 |
| 31-40 yrs | Not Adjusted | 119.83 *** | 17.96 (84.63-155.03) | 0.78 | 32.85 ** | 15.78 (1.92-63.79) | 0.78 |
|  | TVA | 112.31 *** | 17.25 (78.51-146.12) | 0.81 | 26.83 * | 15.05 (-2.68-56.33) | 0.81 |
| 41-50 yrs | Not Adjusted | 71.05 *** | 13.31 (44.97-97.14) | 0.79 | 24.35 ** | 11.68 (1.45-47.25) | 0.79 |
|  | TVA | 68.33 *** | 12.66 (43.52-93.14) | 0.82 | 22.11 ** | 11.04 (0.47-43.74) | 0.82 |
| 51-60 yrs | Not Adjusted | 52.12 *** | 14.51 (23.68-80.55) | 0.78 | 9.86 | 12.73 (-15.09-34.8) | 0.78 |
|  | TVA | 55.73 *** | 13.67 (28.94-82.52) | 0.81 | 11.92 | 11.91 (-11.42-35.27) | 0.81 |
| 61-70 yrs | Not Adjusted | 16.76 ** | 8.51 (0.07-33.44) | 0.79 | -1.44 | 7.46 (-16.06-13.18) | 0.79 |
|  | TVA | 22.22 *** | 7.88 (6.78-37.66) | 0.83 | 1.89 | 6.86 (-11.55-15.34) | 0.83 |
| 71-80 yrs | Not Adjusted | -2.49 | 3.55 (-9.45-4.46) | 0.79 | -2.62 | 3.1 (-8.69-3.45) | 0.79 |
|  | TVA | -1.16 | 3.18 (-7.4-5.07) | 0.84 | -1.55 | 2.76 (-6.96-3.85) | 0.84 |
| 81-90 yrs | Not Adjusted | -7.45 *** | 1.95 (-11.27-3.62) | 0.75 | -6.46 *** | 1.69 (-9.78-3.14) | 0.75 |
|  | TVA | -3.97 ** | 1.8 (-7.49-0.44) | 0.8 | -3.54 ** | 1.55 (-6.58-0.49) | 0.8 |

Notes: Each row represents two coefficients of two DID regression models, stay-at-home order and school closure regression models with raw total population in states, with state and date as fixed effects using Eq. 2. We controlled for COVID-19 confirmed cases to adjust the models using Eq. 4 for the TVA model
\*\*\* $p < 0.01$
\*\* $p < 0.05$
\* $p < 0.1$

**Table C.9:** Effects of lockdown interventions on mental health ED visits of different population groups in counties (weighted by county-level population size per 100, 000 population)

| Group | Model | Stay-at-home Order |  |  | R<sup>2</sup> | School Closure |  |  | R<sup>2</sup> |
| --- | --- | --- | --- | --- | --- | --- | --- | --- | --- |
|  |  | Estimate | SE (95% CI) |  |  | Estimate | SE (95% CI) |  |  |
| Total Population | Not Adjusted | 0.43 *** | 0.04 (0.36-0.5) |  | 0.59 | 0.18 *** | 0.03 (0.13-0.24) |  | 0.59 |
|  | VAT | 1.34×10<sup>-15</sup> ** | 6.56×10<sup>-16</sup> (5.16×10<sup>-17</sup>-2.62×10<sup>-15</sup>) |  | 1 | 1.34×10<sup>-15</sup> ** | 5.31×10<sup>-16</sup> (3.02×10<sup>-16</sup>-2.38×10<sup>-15</sup>) |  | 1 |
| Female | Not Adjusted | 0.3 *** | 0.03 (0.24-0.37) |  | 0.61 | 0.11 *** | 0.03 (0.06-0.17) |  | 0.61 |
|  | VAT | -6.27×10<sup>-16</sup> | 6.12×10<sup>-16</sup> (-1.83×10<sup>-15</sup>-5.72×10<sup>-16</sup>) |  | 1 | -2.22×10<sup>-15</sup> *** | 4.21×10<sup>-16</sup> (-3.05×10<sup>-15</sup>-1.4×10<sup>-15</sup>) |  | 1 |
| Male | Not Adjusted | 0.31 *** | 0.03 (0.25-0.38) |  | 0.61 | 0.1 *** | 0.03 (0.05-0.15) |  | 0.61 |
|  | VAT | 1.96×10<sup>-15</sup> *** | 5.35×10<sup>-16</sup> (9.13×10<sup>-16</sup>-3.01×10<sup>-15</sup>) |  | 1 | -1.76×10<sup>-15</sup> *** | 4.49×10<sup>-16</sup> (-2.64×10<sup>-15</sup>-8.82×10<sup>-16</sup>) |  | 1 |
| Panic disorder | Not Adjusted | 0.17 *** | 0.03 (0.12-0.22) |  | 0.76 | 0.05 ** | 0.02 (0.01-0.09) |  | 0.76 |
|  | VAT | -3.28×10<sup>-15</sup> *** | 3.87×10<sup>-16</sup> (-4.04×10<sup>-15</sup>-2.52×10<sup>-15</sup>) |  | 1 | -8.62×10<sup>-16</sup> *** | 3.04×10<sup>-16</sup> (-1.46×10<sup>-15</sup>-2.66×10<sup>-16</sup>) |  | 1 |
| Reaction to sever stress | Not Adjusted | 0.07 *** | 0.02 (0.03-0.11) |  | 0.86 | 0.04 ** | 0.02 (0.01-0.07) |  | 0.86 |
|  | VAT | 1.84×10<sup>-15</sup> *** | 3.3×10<sup>-16</sup> (1.2×10<sup>-15</sup>-2.49×10<sup>-15</sup>) |  | 1 | 1.17×10<sup>-15</sup> *** | 2.44×10<sup>-16</sup> (6.94×10<sup>-16</sup>-1.65×10<sup>-15</sup>) |  | 1 |
| Major depressive disorder, recurrent | Not Adjusted | 0.13 *** | 0.02 (0.09-0.17) |  | 0.86 | 0.02 | 0.02 (-0.01-0.06) |  | 0.86 |
|  | VAT | -1.13×10<sup>-15</sup> *** | 3.01×10<sup>-16</sup> (-1.72×10<sup>-15</sup>-5.34×10<sup>-16</sup>) |  | 1 | 1.04×10<sup>-15</sup> *** | 2.54×10<sup>-16</sup> (5.42×10<sup>-16</sup>-1.54×10<sup>-15</sup>) |  | 1 |
| Major depressive disorder, single episode | Not Adjusted | 0.12 *** | 0.02 (0.07-0.16) |  | 0.79 | 0.04 ** | 0.02 (0.01-0.08) |  | 0.79 |
|  | VAT | -3.48×10<sup>-16</sup> | 3.67×10<sup>-16</sup> (-1.07×10<sup>-15</sup>-3.72×10<sup>-16</sup>) |  | 1 | 1.9×10<sup>-15</sup> *** | 2.7×10<sup>-16</sup> (1.38×10<sup>-15</sup>-2.43×10<sup>-15</sup>) |  | 1 |
| Attention-deficit hyperactivity | Not Adjusted | 0.03 | 0.02 (-0.01-0.08) |  | 0.89 | -0.04 ** | 0.02 (-0.08-0.01) |  | 0.89 |
|  | VAT | -1.79×10<sup>-15</sup> *** | 3.55×10<sup>-16</sup> (-2.48×10<sup>-15</sup>-1.09×10<sup>-15</sup>) |  | 1 | -6.57×10<sup>-16</sup> ** | 2.71×10<sup>-16</sup> (-1.19×10<sup>-15</sup>-1.25×10<sup>-16</sup>) |  | 1 |
| Insomnia | Not Adjusted | -0.01 | 0.04 (-0.08-0.06) |  | 0.98 | -0.05 * | 0.03 (-0.09-0.0) |  | 0.98 |
|  | VAT | -1.59×10<sup>-15</sup> | 1.68×10<sup>-15</sup> (-4.9×10<sup>-15</sup>-1.71×10<sup>-15</sup>) |  | 1 | 6.64×10<sup>-16</sup> | 1.05×10<sup>-15</sup> (-1.4×10<sup>-15</sup>-2.73×10<sup>-15</sup>) |  | 1 |
| Life management difficulty | Not Adjusted | 0.34 | 0.21 (-0.07-0.76) |  | 0.95 | -0.24 * | 0.14 (-0.51-0.02) |  | 0.95 |
|  | VAT | -7.59×10<sup>-16</sup> | 4.83×10<sup>-15</sup> (-1.02×10<sup>-14</sup>-8.72×10<sup>-15</sup>) |  | 1 | 7.16×10<sup>-16</sup> | 2.85×10<sup>-15</sup> (-4.87×10<sup>-15</sup>-6.3×10<sup>-15</sup>) |  | 1 |
| 81-90 yrs | Not Adjusted | -0.1 *** | 0.03 (-0.17-0.03) |  | 0.9 | 0 | 0.02 (-0.05-0.05) |  | 0.9 |
|  | VAT | -3.09×10<sup>-16</sup> | 5.15×10<sup>-16</sup> (-1.32×10<sup>-15</sup>-7.01×10<sup>-16</sup>) |  | 1 | -1.66×10<sup>-15</sup> *** | 3.64×10<sup>-16</sup> (-2.38×10<sup>-15</sup>-9.51×10<sup>-16</sup>) |  | 1 |
| 71-80 yrs | Not Adjusted | 0 | 0.02 (-0.05-0.04) |  | 0.91 | -0.02 | 0.02 (-0.05-0.01) |  | 0.91 |
|  | VAT | 2.08×10<sup>-15</sup> *** | 3.71×10<sup>-16</sup> (1.36×10<sup>-15</sup>-2.81×10<sup>-15</sup>) |  | 1 | -4.51×10<sup>-16</sup> * | 2.7×10<sup>-16</sup> (-9.8×10<sup>-16</sup>-7.77×10<sup>-17</sup>) |  | 1 |
| 61-70 yrs | Not Adjusted | 0.03 | 0.02 (-0.01-0.08) |  | 0.86 | 4.45×10<sup>-5</sup> | 0.02 (-0.03-0.03) |  | 0.86 |
|  | VAT | -2.46×10<sup>-15</sup> *** | 3.22×10<sup>-16</sup> (-3.09×10<sup>-15</sup>-1.82×10<sup>-15</sup>) |  | 1 | -1.16×10<sup>-16</sup> | 2.29×10<sup>-16</sup> (-5.64×10<sup>-16</sup>-3.32×10<sup>-16</sup>) |  | 1 |
| 51-60 yrs | Not Adjusted | 0.09 *** | 0.02 (0.04-0.13) |  | 0.82 | 0.02 | 0.02 (-0.02-0.05) |  | 0.82 |
|  | VAT | 3.19×10<sup>-16</sup> | 2.97×10<sup>-16</sup> (-2.64×10<sup>-16</sup>-9.02×10<sup>-16</sup>) |  | 1 | -7.71×10<sup>-16</sup> *** | 2.27×10<sup>-16</sup> (-1.22×10<sup>-15</sup>-3.26×10<sup>-16</sup>) |  | 1 |
| 41-50 yrs | Not Adjusted | 0.15 *** | 0.02 (0.11-0.19) |  | 0.81 | 0.02 | 0.02 (-0.01-0.05) |  | 0.81 |
|  | VAT | -2.15×10<sup>-15</sup> *** | 3.24×10<sup>-16</sup> (-2.78×10<sup>-15</sup>-1.52×10<sup>-15</sup>) |  | 1 | 2.2×10<sup>-16</sup> | 2.23×10<sup>-16</sup> (-2.17×10<sup>-16</sup>-6.56×10<sup>-16</sup>) |  | 1 |
| 31-40 yrs | Not Adjusted | 0.18 *** | 0.02 (0.14-0.23) |  | 0.8 | 0.05 *** | 0.02 (0.02-0.09) |  | 0.8 |
|  | VAT | -1.39×10<sup>-15</sup> *** | 3.38×10<sup>-16</sup> (-2.06×10<sup>-15</sup>-7.31×10<sup>-16</sup>) |  | 1 | 3.89×10<sup>-15</sup> *** | 2.73×10<sup>-16</sup> (3.36×10<sup>-15</sup>-4.42×10<sup>-15</sup>) |  | 1 |
| 21-30 yrs | Not Adjusted | 0.09 *** | 0.02 (0.04-0.13) |  | 0.8 | 0.04 ** | 0.02 (0.0-0.07) |  | 0.8 |
|  | VAT | -6.34×10<sup>-16</sup> * | 3.21×10<sup>-16</sup> (-1.26×10<sup>-15</sup>-4.64×10<sup>-18</sup>) |  | 1 | 2.74×10<sup>-15</sup> *** | 2.4×10<sup>-16</sup> (2.27×10<sup>-15</sup>-3.21×10<sup>-15</sup>) |  | 1 |
| 11-20 yrs | Not Adjusted | 0.06 ** | 0.02 (0.01-0.11) |  | 0.81 | 0.03 | 0.02 (-0.01-0.06) |  | 0.81 |
|  | VAT | -1.07×10<sup>-16</sup> | 3.34×10<sup>-16</sup> (-7.61×10<sup>-16</sup>-5.47×10<sup>-16</sup>) |  | 1 | 1.13×10<sup>-15</sup> *** | 2.49×10<sup>-16</sup> (6.43×10<sup>-16</sup>-1.62×10<sup>-15</sup>) |  | 1 |
| 1-10 yrs | Not Adjusted | 0.03 | 0.04 (-0.05-0.11) |  | 0.9 | -0.01 | 0.03 (-0.07-0.04) |  | 0.9 |
|  | VAT | -6.12×10<sup>-16</sup> | 7.07×10<sup>-16</sup> (-2.0×10<sup>-15</sup>-7.74×10<sup>-16</sup>) |  | 1 | 1.49×10<sup>-15</sup> *** | 5.32×10<sup>-16</sup> (4.51×10<sup>-16</sup>-2.53×10<sup>-15</sup>) |  | 1 |

Notes: Each row represents two coefficients of two DID regression models, stay-at-home order and school closure regression models with normalized effects by population size in counties, with county and date as fixed effects using Eq. 1. We controlled for COVID-19 confirmed cases to adjust the models using Eq. 3 for the TVA model
\*\*\* $p < 0.01$
\*\* $p < 0.05$
\* $p < 0.1$

**Table C.10:** Effects of lockdown interventions on mental health ED visits of different population groups in states (weighted by state-level population size)

| Group | Model | Stay-at-home Order |  |  | School Closure |  |  |
| --- | --- | --- | --- | --- | --- | --- | --- |
|  |  | Estimate | SE (95% CI) | R<sup>2</sup> | Estimate | SE (95% CI) | R<sup>2</sup> |
| Total Population | Not Adjusted | 0.29 *** | 0.05 (0.19-0.38) | 0.7 | 0.12 ** | 0.04 (0.03-0.2) | 0.7 |
|  | VAT | 0.32 *** | 0.05 (0.23-0.42) | 0.7 | 0.12 *** | 0.04 (0.04-0.21) | 0.7 |
| Female | Not Adjusted | 0.2 *** | 0.04 (0.12-0.27) | 0.66 | 0.08 ** | 0.03 (0.02-0.15) | 0.66 |
|  | VAT | 0.22 *** | 0.04 (0.15-0.3) | 0.67 | 0.08 ** | 0.03 (0.02-0.15) | 0.67 |
| Male | Not Adjusted | 0.2 *** | 0.04 (0.13-0.28) | 0.67 | 0.07 * | 0.04 (-0.0-0.14) | 0.67 |
|  | VAT | 0.23 *** | 0.04 (0.15-0.31) | 0.68 | 0.07 * | 0.04 (0.0-0.14) | 0.68 |
| Panic disorder | Not Adjusted | 0.1 *** | 0.02 (0.07-0.13) | 0.67 | 0.04 *** | 0.01 (0.02-0.07) | 0.67 |
|  | VAT | 0.09 *** | 0.02 (0.06-0.12) | 0.67 | 0.04 ** | 0.01 (0.01-0.06) | 0.67 |
| Reaction to sever stress | Not Adjusted | 0.02 ** | 0.01 (0.01-0.04) | 0.63 | 0.03 *** | 0.01 (0.02-0.05) | 0.63 |
|  | VAT | 0.02 | 0.01 (-0.0-0.04) | 0.63 | 0.03 *** | 0.01 (0.01-0.04) | 0.63 |
| Major depressive disorder, recurrent | Not Adjusted | 0.06 *** | 0.01 (0.05-0.07) | 0.56 | 0.01 * | 0.0 (-0.0-0.02) | 0.56 |
|  | VAT | 0.06 *** | 0.01 (0.05-0.07) | 0.57 | 0.01 | 0.0 (-0.0-0.01) | 0.57 |
| Major depressive disorder, single episode | Not Adjusted | 0.03 ** | 0.01 (0.0-0.05) | 0.64 | 0.06 *** | 0.01 (0.04-0.08) | 0.64 |
|  | VAT | 0.02 | 0.01 (-0.01-0.05) | 0.65 | 0.06 *** | 0.01 (0.03-0.08) | 0.65 |
| Attention-deficit hyperactivity | Not Adjusted | 0.03 *** | 0.01 (0.02-0.04) | 0.59 | 0 | 0.0 (-0.01-0.01) | 0.58 |
|  | VAT | 0.03 *** | 0.01 (0.02-0.04) | 0.59 | 0 | 0.0 (-0.01-0.0) | 0.59 |
| Insomnia | Not Adjusted | 0 | 0.0 (-0.0-0.0) | 0.69 | 0 | 0.0 (-0.0-0.0) | 0.69 |
|  | VAT | 0 | 0.0 (-0.0-0.0) | 0.71 | 0 | 0.0 (-0.0-0.0) | 0.71 |
| Life management difficulty | Not Adjusted | 0.0 ** | 0.0 (0.0-0.01) | 0.82 | 0 | 0.0 (-0.01-0.0) | 0.82 |
|  | VAT | 0.0 * | 0.0 (-0.0-0.01) | 0.85 | -0.0 * | 0.0 (-0.01--6.17×10<sup>-5</sup>) | 0.85 |
| 81-90 yrs | Not Adjusted | 0 | 0.0 (-0.01-0.0) | 0.5 | 0.01 *** | 0.0 (0.0-0.01) | 0.5 |
|  | VAT | -0.01 ** | 0.0 (-0.01--0.0) | 0.51 | 0.01 ** | 0.0 (0.0-0.01) | 0.51 |
| 71-80 yrs | Not Adjusted | 0.01 ** | 0.0 (0.0-0.02) | 0.56 | 0 | 0.0 (-0.01-0.01) | 0.56 |
|  | VAT | 0 | 0.0 (-0.01-0.01) | 0.57 | 0 | 0.0 (-0.01-0.0) | 0.57 |
| 61-70 yrs | Not Adjusted | 0.02 ** | 0.01 (0.0-0.03) | 0.62 | 0.01 ** | 0.01 (0.0-0.02) | 0.62 |
|  | VAT | 0.01 | 0.01 (-0.0-0.02) | 0.63 | 0.01 | 0.01 (-0.0-0.02) | 0.63 |
| 51-60 yrs | Not Adjusted | 0.06 *** | 0.01 (0.04-0.08) | 0.63 | 0 | 0.01 (-0.02-0.02) | 0.63 |
|  | VAT | 0.05 *** | 0.01 (0.03-0.07) | 0.64 | -0.01 | 0.01 (-0.02-0.01) | 0.64 |
| 41-50 yrs | Not Adjusted | 0.06 *** | 0.01 (0.04-0.08) | 0.61 | 0.03 *** | 0.01 (0.01-0.05) | 0.61 |
|  | VAT | 0.05 *** | 0.01 (0.02-0.07) | 0.62 | 0.02 ** | 0.01 (0.01-0.04) | 0.62 |
| 31-40 yrs | Not Adjusted | 0.06 *** | 0.01 (0.04-0.09) | 0.62 | 0.01 | 0.01 (-0.01-0.03) | 0.62 |
|  | VAT | 0.05 *** | 0.01 (0.03-0.08) | 0.63 | 0 | 0.01 (-0.02-0.02) | 0.63 |
| 21-30 yrs | Not Adjusted | 0.07 *** | 0.01 (0.04-0.09) | 0.63 | 0.03 *** | 0.01 (0.02-0.05) | 0.63 |
|  | VAT | 0.05 *** | 0.01 (0.03-0.08) | 0.64 | 0.03 ** | 0.01 (0.01-0.04) | 0.64 |
| 11-20 yrs | Not Adjusted | 0.04 *** | 0.01 (0.02-0.05) | 0.6 | 0.02 *** | 0.01 (0.01-0.04) | 0.6 |
|  | VAT | 0.03 *** | 0.01 (0.02-0.05) | 0.61 | 0.02 ** | 0.01 (0.01-0.03) | 0.61 |
| 1-10 yrs | Not Adjusted | 0 | 0.0 (-0.0-0.01) | 0.5 | 0 | 0.0 (-0.0-0.0) | 0.5 |
|  | VAT | 0 | 0.0 (-0.01-0.0) | 0.51 | 0 | 0.0 (-0.0-0.0) | 0.51 |

Notes: Each row represents two coefficients of two DID regression models, stay-at-home order and school closure regression models with normalized effects by population size in counties, with county and date as fixed effects using Eq. 1. We controlled for COVID-19 confirmed cases to adjust the models using Eq. 3 for the TVA model
\*\*\* $p < 0.01$
\*\* $p < 0.05$
\* $p < 0.1$

**Table E.11:** Robustness Check for Mental Health Resource Usage

| Criteria | Effect Estimate | SE (95% CI) | R<sup>2</sup> |
| --- | --- | --- | --- |
| Excluding New York and Ohio | $6.2 \times 10^{-5}***$ | $1.2 \times 10^{-5}(3.8 \times 10^{-5} - 8.6 \times 10^{-5})$ | 0.79 |
| Including all 2019 data | $9.4 \times 10^{-5}***$ | $8.6 \times 10^{-5}(7.7 \times 10^{-5} - 1.1 \times 10^{-4})$ | 0.79 |

**Figure D.5:**
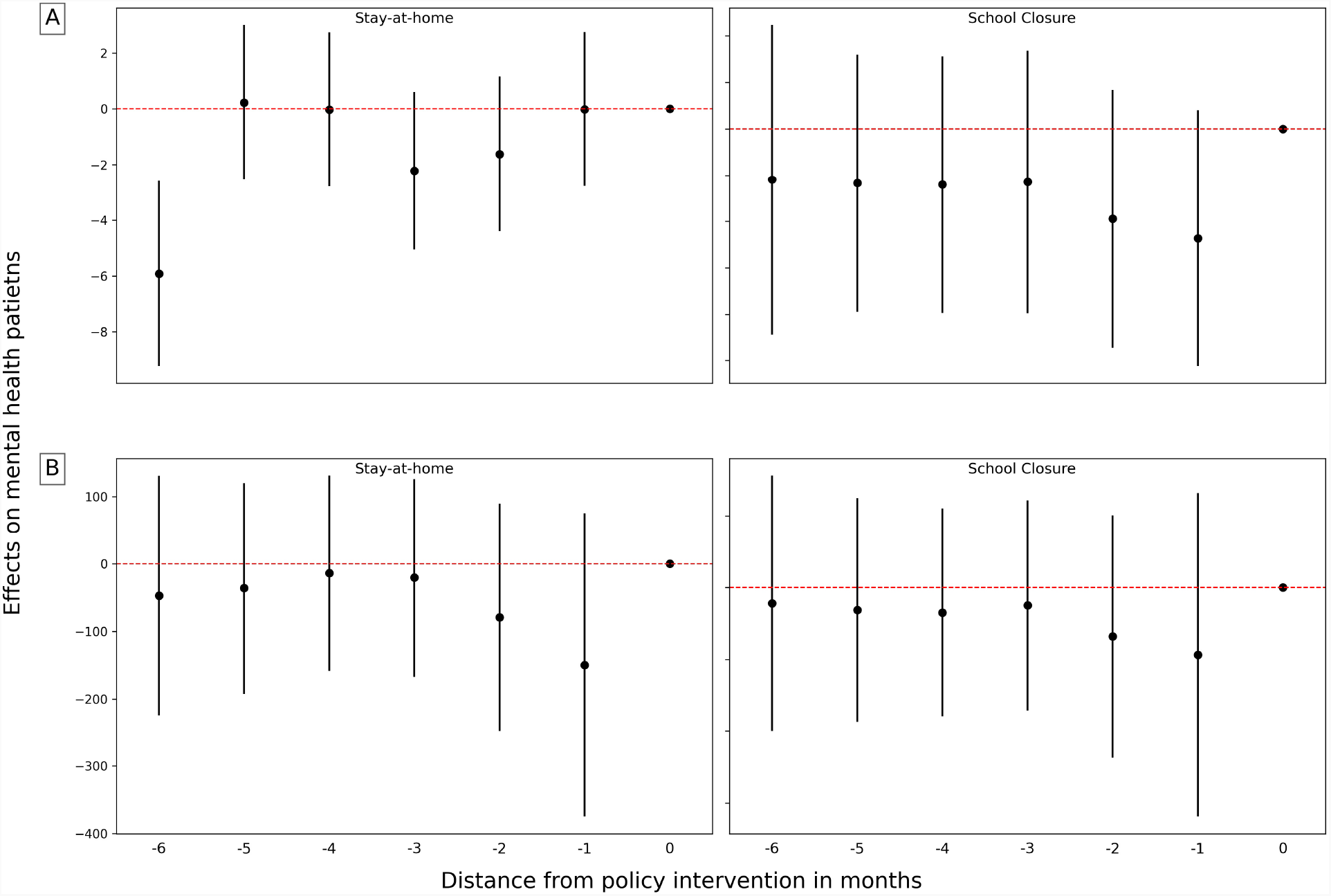
Event study of lockdowns effect (stay-at-home and school closure) in counties (A) and states (B) using 6-months pre-policy patients numbers as counterfactual. Coefficients are shown with their corresponding 95% confidence interval.

**Table E.12:** Robustness Check for Mental Health ED Visits

| Criteria | Effect Estimate | SE (95% CI) | $R^2$ |
| --- | --- | --- | --- |
| Excluding New York and Ohio | $0.22 \times 10^{-5}***$ | $0.06 \times 10^{-5}(0.09 \times 10^{-5} - 0.3 \times 10^{-5})$ | 0.67 |
| Including all 2019 data | $0.35 \times 10^{-5}***$ | $0.04 \times 10^{-5}(0.27 \times 10^{-5} - 0.44 \times 10^{-4})$ | 0.70 |

**Table F.13:** Effects on mental health in counties *k*-months after lockdown

| Group | Period | Stay-at-home Order |  |  | School Closure |  |  |
| --- | --- | --- | --- | --- | --- | --- | --- |
|  |  | Estimat | SE (95% CI) | R<sup>2</sup> | Estimate | SE (95% CI) | R<sup>2</sup> |
| Total Population | 1-month | 5.4534 *** | 0.65 (1.39-3.923) | 2.729 | 1.63 | 1.1 (-0.53-3.79) | 0.73 |
|  | 5-month | 8.1074 *** | 1.41 (1.203-6.741) | 5.75 | 2.22 ** | 0.81 (0.64-3.8) | 0.73 |
|  | 9-month | 9.79 *** | 0.84 (8.15-11.43) | 0.73 | 2.35 *** | 0.74 (0.89-3.81) | 0.73 |
| Female | 1-month | 3.95 *** | 0.62 (2.73-5.17) | 0.74 | 1.38 ** | 0.61 (0.18-2.57) | 0.73 |
|  | 5-month | 6.08 *** | 0.49 (5.12-7.05) | 0.74 | 2.03 *** | 0.45 (1.14-2.92) | 0.73 |
|  | 9-month | 7.32 *** | 0.48 (6.38-8.26) | 0.73 | 2.21 *** | 0.42 (1.38-3.03) | 0.73 |
| Male | 1-month | 2.25 *** | 0.61 (1.05-3.44) | 0.73 | 4.46 *** | 0.56 (3.37-5.55) | 0.73 |
|  | 5-month | 3.05 *** | 0.47 (2.13-3.96) | 0.73 | 3.81 *** | 0.47 (2.89-4.73) | 0.73 |
|  | 9-month | 3.68 *** | 0.44 (2.81-4.55) | 0.73 | 0.04 | 0.39 (-0.72-0.81) | 0.73 |
| Panic disorder | 1-month | 1.71 *** | 0.29 (1.13-2.28) | 0.75 | 0.64 ** | 0.29 (0.08-1.2) | 0.74 |
|  | 5-month | 3.34 *** | 0.24 (2.87-3.8) | 0.75 | 1.12 *** | 0.21 (0.7-1.54) | 0.74 |
|  | 9-month | 4.06 *** | 0.23 (3.6-4.52) | 0.74 | 1.25 *** | 0.2 (0.84-1.65) | 0.74 |
| Reaction to sever stress | 1-month | 2.26 *** | 0.3 (1.68-2.84) | 0.71 | 0.3 | 0.31 (-0.3-0.91) | 0.69 |
|  | 5-month | 2.93 *** | 0.23 (2.47-3.39) | 0.7 | 0.69 *** | 0.22 (0.26-1.12) | 0.7 |
|  | 9-month | 3.38 *** | 0.23 (2.93-3.83) | 0.7 | 0.65 *** | 0.2 (0.26-1.05) | 0.7 |
| Major depressive disorder, recurrent | 1-month | 0.85 *** | 0.24 (0.37-1.32) | 0.72 | -0.05 | 0.23 (-0.5-0.39) | 0.71 |
|  | 5-month | 1.81 *** | 0.19 (1.43-2.18) | 0.72 | 0.35 ** | 0.17 (0.01-0.69) | 0.71 |
|  | 9-month | 2.27 *** | 0.19 (1.91-2.63) | 0.71 | 0.36 ** | 0.16 (0.04-0.68) | 0.71 |
| Major depressive disorder, single episode | 1-month | 0.13 | 0.17 (-0.2-0.47) | 0.72 | -0.21 | 0.16 (-0.52-0.1) | 0.71 |
|  | 5-month | 0.69 *** | 0.13 (0.43-0.95) | 0.71 | 0.03 | 0.12 (-0.2-0.26) | 0.71 |
|  | 9-month | 0.96 *** | 0.13 (0.71-1.21) | 0.71 | 0.09 | 0.11 (-0.12-0.31) | 0.71 |
| Attention-deficit hyperactivity | 1-month | 1.43 *** | 0.23 (0.97-1.89) | 0.62 | 0.25 | 0.22 (-0.18-0.67) | 0.62 |
|  | 5-month | 1.56 *** | 0.18 (1.21-1.91) | 0.62 | 0.3 * | 0.16 (-0.01-0.61) | 0.62 |
|  | 9-month | 1.78 *** | 0.17 (1.45-2.11) | 0.62 | 0.36 ** | 0.14 (0.08-0.64) | 0.62 |
| Insomnia | 1-month | 0.2 *** | 0.06 (0.08-0.32) | 0.58 | 0.05 | 0.05 (-0.05-0.16) | 0.56 |
|  | 5-month | 0.22 *** | 0.05 (0.12-0.31) | 0.57 | 0.06 | 0.04 (-0.02-0.14) | 0.57 |
|  | 9-month | 0.27 *** | 0.05 (0.17-0.35) | 0.57 | 0.03 | 0.04 (-0.05-0.1) | 0.57 |
| Life management difficulty | 1-month | -0.34 | 0.77 (-1.84-1.17) | 0.3 | -1.98 ** | 0.96 (-3.87-0.1) | 0.32 |
|  | 5-month | 0.88 | 0.7 (-0.49-2.24) | 0.32 | -3.66 *** | 0.6 (-4.83-2.48) | 0.31 |
|  | 9-month | 1.34 * | 0.78 (-0.18-2.86) | 0.3 | -5.14 *** | 0.5 (-6.12-4.17) | 0.3 |
| 1-10 yrs | 1-month | 0.25 | 0.29 (-0.32-0.82) | 0.61 | -0.45 * | 0.27 (-0.97-0.07) | 0.61 |
|  | 5-month | 0.69 *** | 0.22 (0.27-1.12) | 0.61 | -0.19 | 0.19 (-0.56-0.19) | 0.61 |
|  | 9-month | 1.09 *** | 0.2 (0.69-1.49) | 0.61 | 0.17 | 0.18 (-0.17-0.51) | 0.61 |
| 11-20 yrs | 1-month | 1.54 *** | 0.34 (0.87-2.22) | 0.62 | 0.3 | 0.33 (-0.35-0.95) | 0.62 |
|  | 5-month | 1.68 *** | 0.26 (1.16-2.19) | 0.62 | 0.4 | 0.24 (-0.08-0.87) | 0.62 |
|  | 9-month | 2.08 *** | 0.25 (1.59-2.57) | 0.62 | 0.6 *** | 0.22 (0.17-1.02) | 0.62 |
| 21-30 yrs | 1-month | 1.8 *** | 0.24 (1.32-2.27) | 0.76 | 0.37 | 0.25 (-0.11-0.86) | 0.75 |
|  | 5-month | 2.75 *** | 0.2 (2.37-3.14) | 0.76 | 0.65 *** | 0.18 (0.3-1.0) | 0.75 |
|  | 9-month | 3.39 *** | 0.19 (3.02-3.77) | 0.75 | 0.78 *** | 0.17 (0.46-1.11) | 0.75 |
| 31-40 yrs | 1-month | 2.55 *** | 0.27 (2.02-3.08) | 0.75 | 0.76 *** | 0.27 (0.23-1.29) | 0.75 |
|  | 5-month | 3.11 *** | 0.21 (2.69-3.53) | 0.75 | 0.9 *** | 0.19 (0.52-1.28) | 0.75 |
|  | 9-month | 3.52 *** | 0.21 (3.12-3.92) | 0.75 | 0.88 *** | 0.18 (0.53-1.23) | 0.75 |
| 41-50 yrs | 1-month | 1.62 *** | 0.21 (1.2-2.04) | 0.74 | 0.69 *** | 0.21 (0.28-1.09) | 0.74 |
|  | 5-month | 2.02 *** | 0.17 (1.7-2.35) | 0.74 | 0.82 *** | 0.15 (0.52-1.11) | 0.74 |
|  | 9-month | 2.21 *** | 0.16 (1.9-2.52) | 0.74 | 0.72 *** | 0.14 (0.45-0.99) | 0.74 |
| 51-60 yrs | 1-month | 0.85 *** | 0.26 (0.34-1.36) | 0.72 | 0.03 | 0.24 (-0.43-0.5) | 0.72 |
|  | 5-month | 1.4 *** | 0.2 (1.01-1.79) | 0.73 | 0.3 * | 0.18 (-0.04-0.65) | 0.73 |
|  | 9-month | 1.62 *** | 0.19 (1.25-1.99) | 0.73 | 0.2 | 0.16 (-0.11-0.52) | 0.73 |
| 61-70 yrs | 1-month | -0.08 | 0.18 (-0.44-0.27) | 0.73 | -0.1 | 0.16 (-0.41-0.21) | 0.73 |
|  | 5-month | 0.41 *** | 0.14 (0.14-0.68) | 0.73 | 0 | 0.12 (-0.23-0.23) | 0.73 |
|  | 9-month | 0.57 *** | 0.13 (0.32-0.82) | 0.73 | -0.11 | 0.11 (-0.32-0.1) | 0.73 |
| 71-80 yrs | 1-month | -0.27 ** | 0.1 (-0.46-0.08) | 0.73 | -0.05 | 0.08 (-0.2-0.11) | 0.73 |
|  | 5-month | -0.01 | 0.07 (-0.15-0.13) | 0.73 | -0.04 | 0.06 (-0.15-0.08) | 0.73 |
|  | 9-month | 0.04 | 0.07 (-0.09-0.17) | 0.73 | -0.07 | 0.05 (-0.17-0.04) | 0.73 |
| 81-90 yrs | 1-month | -0.14 *** | -0.04 (-0.06-0.21) | 0.68 | -0.24 *** | 0.06 (-0.36-0.12) | 0.68 |
|  | 5-month | -0.13 *** | -0.03 (-0.07-0.19) | 0.68 | -0.23 *** | 0.04 (-0.32-0.15) | 0.68 |
|  | 9-month | -0.24 *** | 0.05 (-0.34-0.14) | 0.68 | -0.25 *** | 0.04 (-0.33-0.17) | 0.68 |

Notes: Each row represents two coefficients of two DID regression models, stay-at-home order and school closure regression models with raw total population in counties, with county and date as fixed effects using Eq. 1. Each mental health group has been evaluated on 3 different points of time $z$ with regard to $k=\{1,5,9\}$ -months after lockdown, to observe the dynamic effect of lockdown.
\*\*\* $p < 0.01$
\*\* $p < 0.05$
\* $p < 0.1$

**Table F.14:** Effects on mental health in states *z*-months after lockdown

| Group | Period | Stay-at-home Order |  |  | School Closure |  |  |
| --- | --- | --- | --- | --- | --- | --- | --- |
|  |  | Estimate | SE (95% CI) | R<sup>2</sup> | Estimate | SE (95% CI) | R<sup>2</sup> |
| Total Population | 1-month | 274.81 * | 139.95 (0.51-549.11) | 0.78 | 35.86 | 126.85 (-212.76-284.48) | 0.78 |
|  | 5-month | 384.0 *** | 108.19 (171.95-596.05) | 0.78 | 96.43 | 96.23 (-92.17-285.04) | 0.78 |
|  | 9-month | 466.62 *** | 102.86 (265.01-668.23) | 0.78 | 117.18 | 90.38 (-59.95-294.32) | 0.78 |
| Female | 1-month | 181.37 ** | 72.74 (38.79-323.95) | 0.79 | 48.47 | 65.99 (-80.87-177.81) | 0.78 |
|  | 5-month | 266.64 *** | 57.24 (154.44-378.83) | 0.79 | 97.35 * | 50.92 (-2.45-197.14) | 0.78 |
|  | 9-month | 322.89 *** | 55.02 (215.06-430.73) | 0.79 | 113.61 ** | 48.35 (18.85-208.38) | 0.79 |
| Male | 1-month | 93.85 | 67.37 (-38.19-225.89) | 0.78 | -26.18 | 60.88 (-145.5-93.14) | 0.77 |
|  | 5-month | 117.9 ** | 51.19 (17.57-218.23) | 0.78 | -16.06 | 45.49 (-105.22-73.09) | 0.77 |
|  | 9-month | 144.06 *** | 48.1 (49.78-238.34) | 0.78 | -11.9 | 42.25 (-94.72-70.91) | 0.78 |
| Panic disorder | 1-month | 79.53 ** | 29.55 (21.61-137.45) | 0.8 | 30.78 | 27.17 (-22.47-84.03) | 0.8 |
|  | 5-month | 140.9 *** | 24.04 (93.78-188.02) | 0.8 | 62.69 *** | 21.44 (20.66-104.72) | 0.8 |
|  | 9-month | 170.79 *** | 23.48 (124.76-216.81) | 0.8 | 71.45 *** | 20.64 (31.0-111.9) | 0.79 |
| Reaction to sever stress | 1-month | 86.29 *** | 24.49 (38.28-134.3) | 0.78 | 16.74 | 23.18 (-28.7-62.18) | 0.76 |
|  | 5-month | 107.24 *** | 19.45 (69.12-145.35) | 0.77 | 34.65 * | 17.57 (0.2-69.09) | 0.77 |
|  | 9-month | 125.04 *** | 18.98 (87.85-162.24) | 0.77 | 36.31 ** | 16.68 (3.62-68.99) | 0.77 |
| Major depressive disorder, recurrent | 1-month | 38.17 * | 20.02 (-1.08-77.41) | 0.78 | 1.84 | 18.05 (-33.54-37.22) | 0.78 |
|  | 5-month | 70.4 *** | 15.98 (39.08-101.72) | 0.78 | 21.61 | 14.18 (-6.18-49.39) | 0.78 |
|  | 9-month | 88.15 *** | 15.42 (57.93-118.36) | 0.78 | 24.53 * | 13.54 (-2.02-51.07) | 0.78 |
| Major depressive disorder, single episode | 1-month | 8.59 | 13.37 (-17.62-34.79) | 0.78 | -4.7 | 12.14 (-28.5-19.1) | 0.78 |
|  | 5-month | 26.49 ** | 10.47 (5.96-47.01) | 0.78 | 5.28 | 9.31 (-12.98-23.53) | 0.78 |
|  | 9-month | 35.63 *** | 10.04 (15.95-55.32) | 0.78 | 8.82 | 8.82 (-8.46-26.1) | 0.78 |
| Attention-deficit hyperactivity | 1-month | 49.23 ** | 17.55 (14.83-83.63) | 0.74 | 10.04 | 15.75 (-20.83-40.9) | 0.74 |
|  | 5-month | 51.22 *** | 13.27 (25.2-77.23) | 0.74 | 11.71 | 11.77 (-11.36-34.78) | 0.74 |
|  | 9-month | 59.13 *** | 12.48 (34.67-83.59) | 0.74 | 15.16 | 10.93 (-6.27-36.59) | 0.74 |
| Insomnia | 1-month | 3.62 ** | 1.64 (0.42-6.83) | 0.76 | 1.96 | 1.46 (-0.9-4.82) | 0.75 |
|  | 5-month | 5.03 *** | 1.33 (2.42-7.65) | 0.76 | 2.89 ** | 1.16 (0.61-5.17) | 0.75 |
|  | 9-month | 5.83 *** | 1.27 (3.34-8.33) | 0.76 | 2.32 ** | 1.09 (0.18-4.47) | 0.76 |
| Life management difficulty | 1-month | 0.2 | 2.51 (-4.71-5.11) | 0.45 | -4.38 | 3.51 (-11.26-2.49) | 0.48 |
|  | 5-month | 4.51 * | 2.4 (-0.19-9.2) | 0.44 | -9.5 *** | 2.3 (-14.01-4.98) | 0.46 |
|  | 9-month | 6.51 ** | 2.55 (1.51-11.51) | 0.43 | -14.35 *** | 1.96 (-18.19-10.51) | 0.43 |
| 1-10 yrs | 1-month | 3.05 | 19.82 (-35.79-41.89) | 0.71 | -16.58 | 17.64 (-51.15-17.98) | 0.71 |
|  | 5-month | 16.32 | 14.84 (-12.77-45.42) | 0.71 | -4.53 | 13.09 (-30.18-21.12) | 0.71 |
|  | 9-month | 29.48 ** | 14.12 (1.79-57.16) | 0.72 | 9 | 12.37 (-15.24-33.25) | 0.72 |
| 11-20 yrs | 1-month | 67.83 ** | 29.37 (10.26-125.4) | 0.74 | 11.1 | 26.53 (-40.9-63.11) | 0.73 |
|  | 5-month | 67.56 *** | 22.32 (23.81-111.32) | 0.73 | 16.59 | 19.87 (-22.36-55.54) | 0.73 |
|  | 9-month | 82.17 *** | 21.12 (40.77-123.57) | 0.73 | 27.29 | 18.55 (-9.06-63.65) | 0.73 |
| 21-30 yrs | 1-month | 63.26 *** | 19.86 (24.33-102.19) | 0.8 | 10.85 | 18.46 (-25.33-47.02) | 0.79 |
|  | 5-month | 90.9 *** | 15.97 (59.6-122.19) | 0.79 | 23.01 | 14.3 (-5.02-51.03) | 0.79 |
|  | 9-month | 112.63 *** | 15.57 (82.11-143.14) | 0.79 | 28.11 ** | 13.68 (1.29-54.92) | 0.79 |
| 31-40 yrs | 1-month | 89.59 *** | 24.18 (42.19-136.99) | 0.78 | 26.6 | 22.09 (-16.7-69.91) | 0.78 |
|  | 5-month | 105.21 *** | 18.86 (68.24-142.17) | 0.78 | 32.31 * | 16.81 (-0.64-65.25) | 0.78 |
|  | 9-month | 119.83 *** | 17.96 (84.63-155.03) | 0.78 | 32.85 ** | 15.78 (1.92-63.79) | 0.78 |
| 41-50 yrs | 1-month | 52.44 *** | 18.13 (16.9-87.98) | 0.79 | 22.19 | 16.44 (-10.02-54.41) | 0.78 |
|  | 5-month | 63.72 *** | 14.08 (36.12-91.32) | 0.78 | 27.21 ** | 12.52 (2.67-51.74) | 0.78 |
|  | 9-month | 71.05 *** | 13.31 (44.97-97.13) | 0.79 | 24.35 ** | 11.68 (1.45-47.25) | 0.79 |
| 51-60 yrs | 1-month | 28.05 | 20.03 (-11.22-67.31) | 0.78 | 3.68 | 17.93 (-31.47-38.83) | 0.77 |
|  | 5-month | 45.37 *** | 15.47 (15.04-75.7) | 0.78 | 13 | 13.69 (-13.83-39.82) | 0.77 |
|  | 9-month | 52.12 *** | 14.51 (23.68-80.55) | 0.78 | 9.86 | 12.73 (-15.09-34.8) | 0.78 |
| 61-70 yrs | 1-month | -3.42 | 11.83 (-26.6-19.76) | 0.79 | -5.24 | 10.58 (-25.98-15.51) | 0.78 |
|  | 5-month | 12.32 | 9.11 (-5.53-30.17) | 0.79 | 1.44 | 8.05 (-14.33-17.21) | 0.78 |
|  | 9-month | 16.76 * | 8.51 (0.07-33.44) | 0.79 | -1.44 | 7.46 (-16.06-13.18) | 0.79 |
| 71-80 yrs | 1-month | -11.02 ** | 5.02 (-20.86-1.19) | 0.79 | -3.96 | 4.41 (-12.59-4.68) | 0.79 |
|  | 5-month | -3.76 | 3.83 (-11.26-3.74) | 0.79 | -1.88 | 3.34 (-8.44-4.67) | 0.79 |
|  | 9-month | -2.49 | 3.55 (-9.45-4.47) | 0.79 | -2.62 | 3.1 (-8.69-3.45) | 0.79 |
| 81-90 yrs | 1-month | -9.6 *** | 2.92 (-15.31-3.88) | 0.75 | -6.67 ** | 2.53 (-11.62-1.72) | 0.75 |
|  | 5-month | -7.74 *** | 2.15 (-11.94-3.53) | 0.75 | -6.01 *** | 1.87 (-9.67-2.35) | 0.75 |
|  | 9-month | -7.45 *** | 1.95 (-11.27-3.62) | 0.75 | -6.46 *** | 1.69 (-9.78-3.14) | 0.75 |

Notes: Each row represents two coefficients of two DID regression models, stay-at-home order and school closure regression models with raw total population in states, with state and date as fixed effects using Eq. 2. Each mental health group has been evaluated on 3 different points of time $z$ with regard to $z=\{1,5,9\}$ -months after lockdown, to observe the dynamic effect of lockdown.
\*\*\* $p < 0.01$
\*\* $p < 0.05$
\* $p < 0.1$
\*\*\* $p < 0.01$ \*\* $p < 0.05$ \* $p < 0.1$

**Figure G.6:**
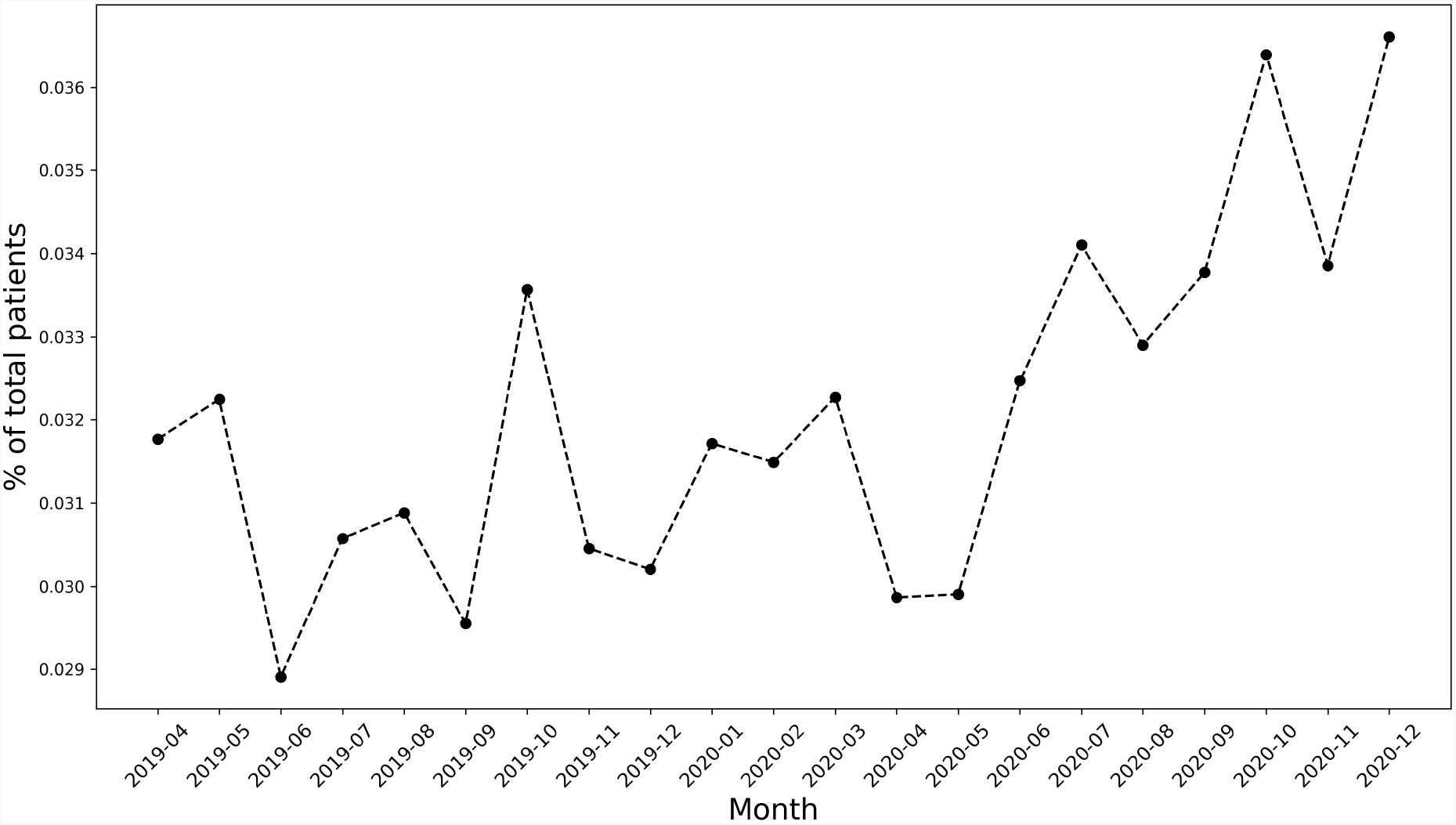
Mental health patients over time

**Figure G.7:**
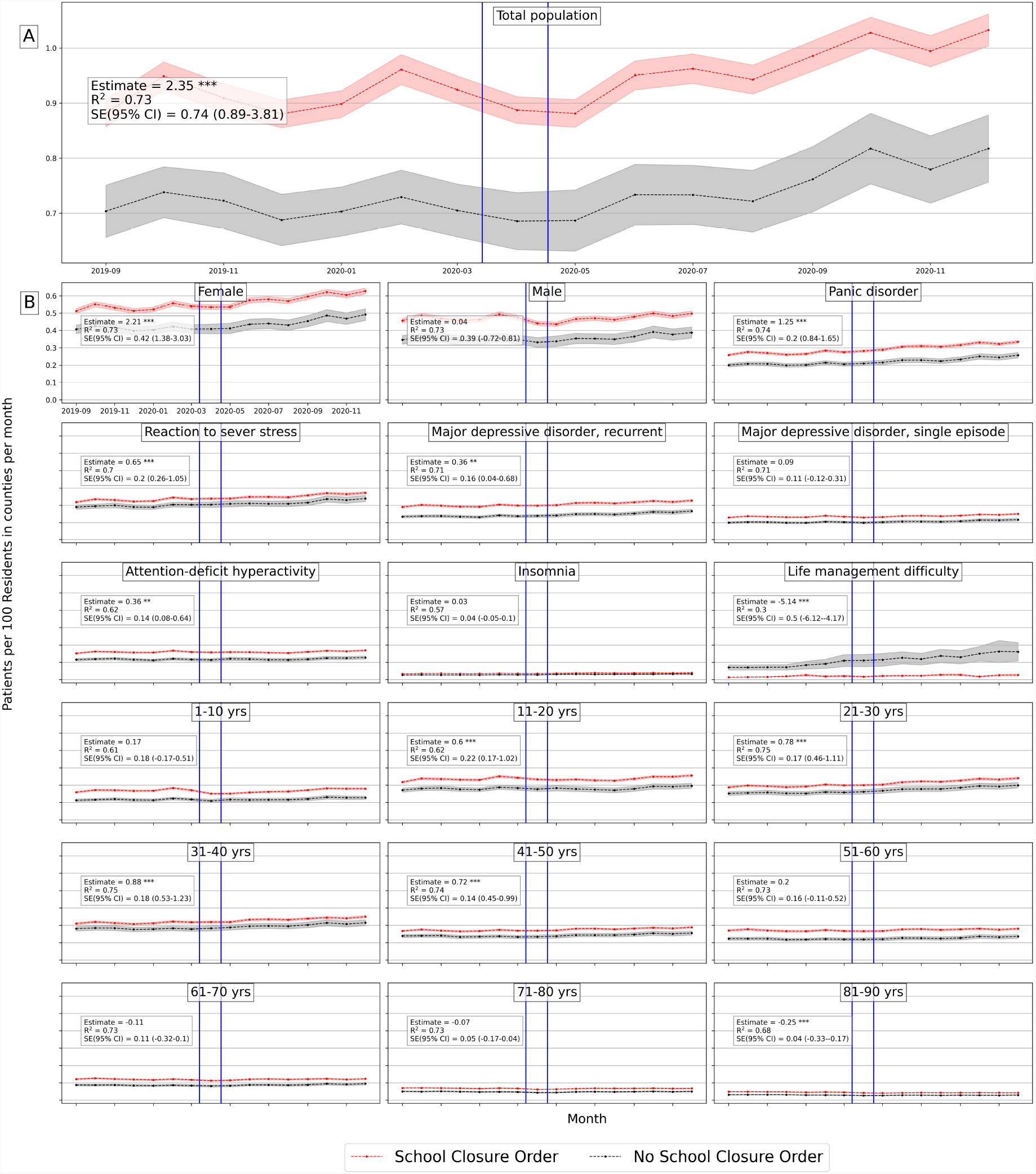
Average number of mental health patients over time (September 2019 - December 2020) in counties with school closure orders and without. Vertical lines show the first school closure on 3/10/2020 and last on 4/28/2020 across United States. Difference-in-differences estimates are included for each population group. (Detailed average percentage changes are listed in Table B.3) *\* * * p <* 0.01 *\* * p <* 0.05 *\* p <* 0.1

**Figure G.8:**
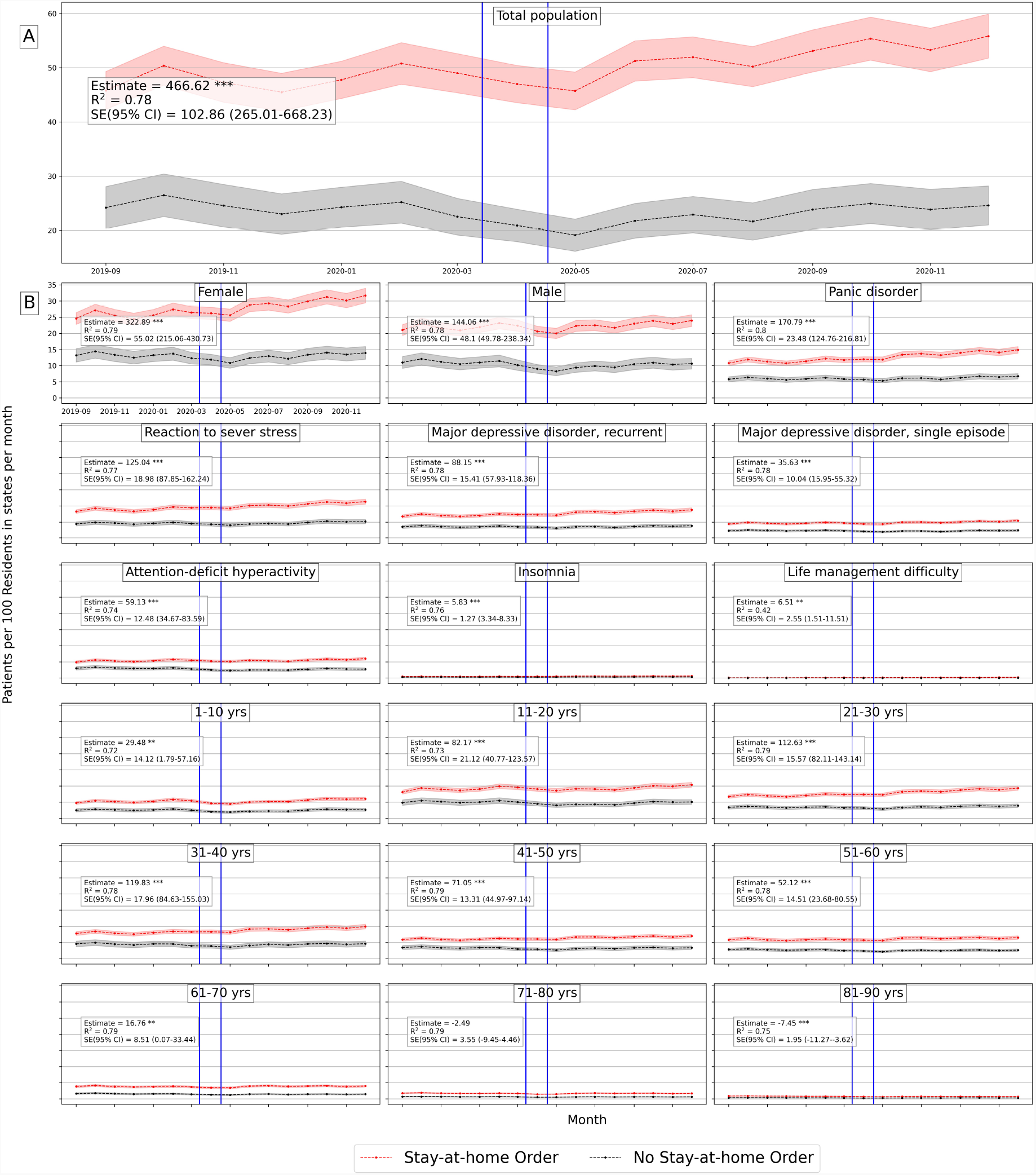
Average number of mental health patients over time (September 2019 - December 2020) in states with stay-at-home orders and without. Vertical lines show the first stay-at-home order on 3/14/2020 and last 0n 4/07/2020 across United States. Difference-in-differences estimates are included for each population group. (Detailed average percentage changes are listed in Table B.4) *\* * * p <* 0.01 *\* * p <* 0.05 *\* p <* 0.1

**Figure G.9:**
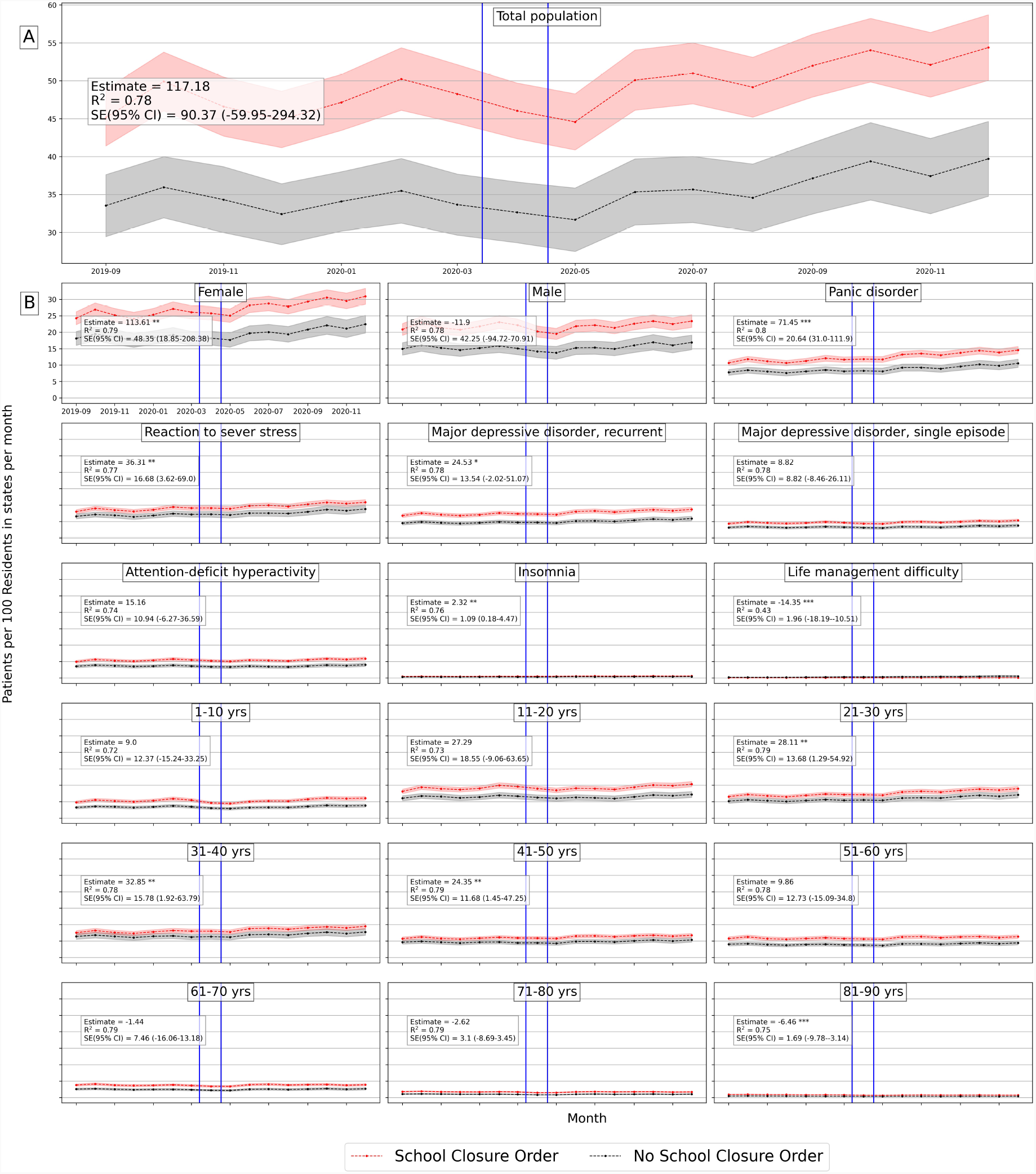
Average number of mental health patients over time (September 2019 - December 2020) in states with school closure orders and without. Vertical lines show the first school closure on 3/10/2020 and last on 4/28/2020 across United States. Difference-in-differences estimates are included for each population group. (Detailed average percentage changes are listed in Table B.4) *\* * * p <* 0.01 *\* * p <* 0.05 *\* p <* 0.1

**Figure G.10:**
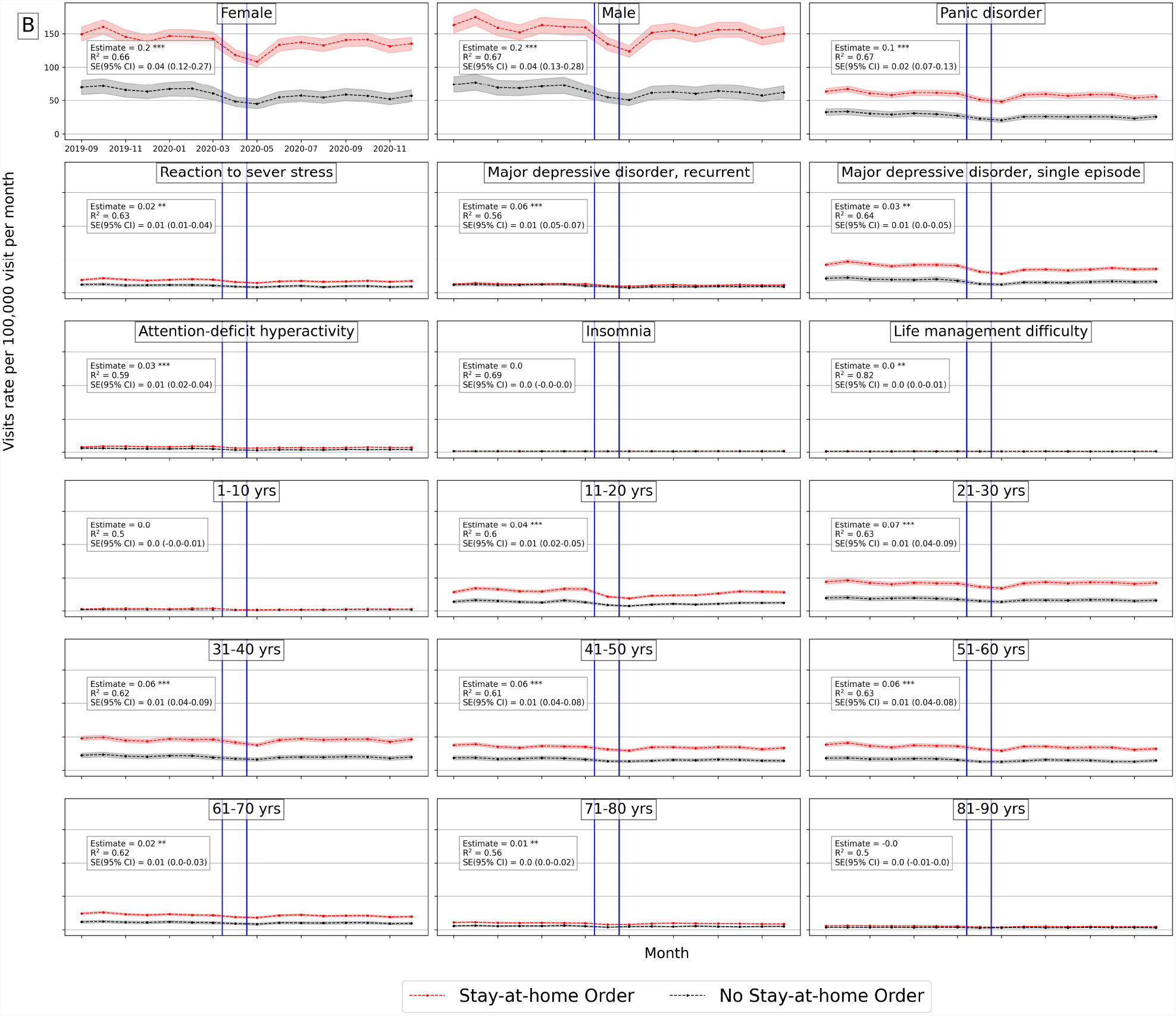
Average number mental health ED visits over time (September 2019 - December 2020) in counties with stay-at-home orders and without. Vertical lines show the first stay-at-home order on 3/14/2020 and last 0n 4/07/2020 across United States. Difference-in-differences estimates are included for each population *\* * * p <* 0.01 *\* * p <* 0.05 *\* p <* 0.1

**Figure G.11:**
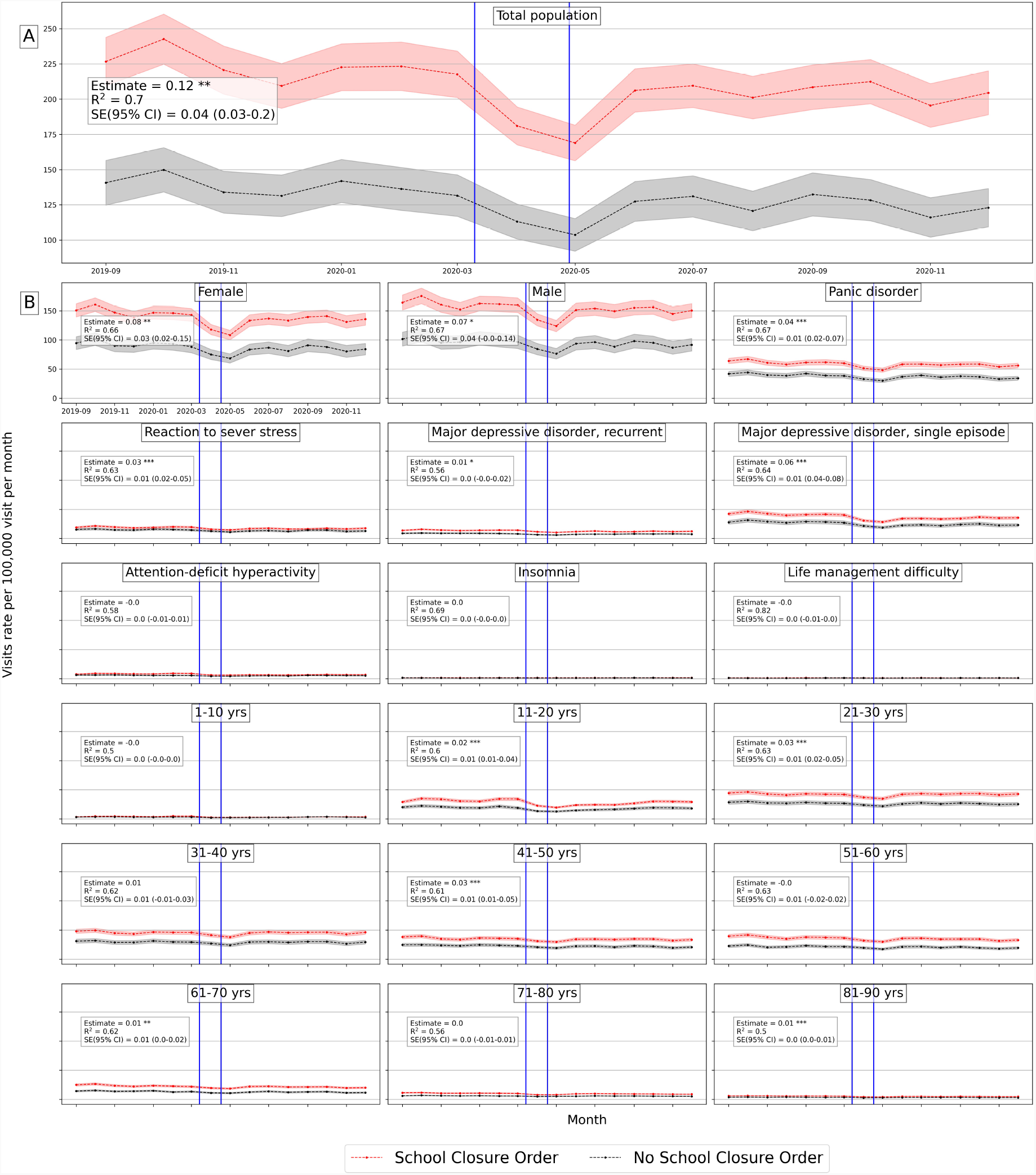
Average number of mental health ED visits over time (September 2019 - December 2020) in counties with school closure orders and without. Vertical lines show the first school closure on 3/10/2020 and last on 4/28/2020 across United States. Difference-in-differences estimates are included for each population group *\* * * p <* 0.01 *\* * p <* 0.05 *\* p <* 0.1

## Footnotes

1 U.S. Census Bureau December Survey https://www.census.gov/data/tables/2020/demo/hhp/hhp21.html

2 COVIDVis https://covidvis.berkeley.edu/#lockdown_section

## References

[1] Alison Abbott. COVID’s mental-health toll: How scientists are tracking a surge in depression. Nature, 590(7845), February 2021.

[2] Abi Adams-Prassl, Teodora Boneva, Marta Golin, and Christopher Rauh. The impact of the coronavirus lockdown on mental health: Evidence from the US. Cambridge Working Papers in Economics 2037, May 2020.

[3] Roy M. Anderson, Hans Heesterbeek, Don Klinkenberg, and T. Déirdre Hollingsworth. How will country-based mitigation measures influence the course of the COVID-19 epidemic? The Lancet, 395(10228):931–934, March 2020.

[4] Annette Bauera, Martin Knappa, and Michael Parsonage. Lifetime costs of perinatal anxiety and depression. Journal of Affective Disorders, 192(1):83–90, March 2016.

[5] Lisa F. Berkman and S. Leonard Syme. Social networks, host resistance, and mortality: A nine-year follow-up study of Alameda County residents. American Journal of Epidemiology, 109(2):186–204, 1979.

[6] D.E. Bloom, E.T. Cafiero, E. Jané-Llopis, S. Abrahams-Gessel, et al. The global economic burden of noncommunicable diseases. Geneva. Technical report, World Economic Forum, 2011.

[7] Abel Brodeur, Andrew E. Clark, Sarah Fleched, and Nattavudh Powdthavee. COVID-19, lockdowns and well-being: Evidence from Google Trends. Journal of Public Economics, 193:104346, January 2021.

[8] Samantha K. Brooks, Rebecca K. Webster, Louise E. Smith, Lisa Woodlan, Simon Wessely, Neil Greenberg, and Gideon James Rubin. The psychological impact of quarantine and how to reduce it: Rapid review of the evidence. The Lancent, 395(10227):912–920, March 2020.

[9] Melisa Bubonya, Deborah A. Cobb-Clark, and Mark Wooden. Mental Health and Productivity at Work: Does What You Do Matter? IZA Discussion Papers 9879, Institute of Labor Economics (IZA), 2016. http://hdl.handle.net/10419/141638.

[10] Vera Clemens, Peter Deschamps, Jörg M. Fegert, Dimitris Anagnostopoulos, Sue Bailey, Maeve Doyle, Stephan Eliez, Anna Sofie Hansen, Johannes Hebebrand, Manon Hillegers, Brian Jacobs, Andreas Karwautz, Eniko Kiss, Konstantinos Kotsis, Hojka Gregoric Kumperscak, Milica Pejovic-Milovancevic, Anne Marie Røaberg Christensen, Jean-Philippe Raynaud, Hannu Westerinen, and Piret Visnapuu-Bernadt. Potential efects of “social” distancing measures and school lockdown on child and adolescent mental health. European Child & Adolescent Psychiatry, 29(6):739–742, June 2020.

[11] Francesca Cornaglia, Elena Crivellaro, and Sandra McNally. Mental health and education decisions. Labour Economics, 33:1–12, 2015.

[12] Domenico Cucinotta and Vanelli Maurizio. WHO declares COVID-19 a pandemic. Acta bio-medica, 91(1):157–160, March 2020.

[13] W. Cullen, G. Gulati, and B. D. Kelly. Mental health in the COVID-19 pandemic. QJM: An International Journal of Medicine, 113(5):311–312, May 2020.

[14] Mark É. Czeisler, Rashon I. Lane, Emiko Petrosky, Joshua F. Wiley, Aleta Christensen, Rashid Njai, Matthew D. Weaver, Rebecca Robbins, Elise R. Facer-Childs, Laura K. Barger, Charles A. Czeisler, Mark E. Howard, and Shantha M. W. Rajaratnam. Mental health, substance use, and suicidal ideation during the COVID-19 pandemic — United States, June 24–30, 2020. Morbidity and Mortality Weekly Report 32, Centers for Disease Control and Prevention (CDC), August 2020.

[15] Srikant Devaraj and Pankaj C. Patel. Change in psychological distress in response to changes in reduced mobility during the early 2020 COVID-19 pandemic: Evidence of modest effects from the U.S. Social Science & Medicine, 270:113615, February 2021.

[16] Lu Dong and Jennifer Bouey. Public mental health crisis during COVID-19 pandemic, China. Emerging Infectious Diseases, 26(7):1616–1618, July 2020.

[17] Timon Elmer, Kieran Mepham, and Christoph Stadtfeld. Students under lockdown: Comparisons of students’ social networks and mental health before and during the COVID-19 crisis in Switzerland. PLoS ONE, 15(7):e0236337, July 2020.

[18] Marta Fana, Sergio Torrejón Pérez, and Enrique Fernández-Macías. Employment impact of Covid-19 crisis: from short term effects to long terms prospects. Journal of Industrial and Business Economic, 47:391–410, July 2020.

[19] Bita Fayaz Farkhad and Dolores Albarracín. Insights on the implications of COVID-19 mitigation measures for mental health. Economics & Human Biology, 40:100963, January 2021.

[20] Massimiliano Ferraresi, Christos Kotsogiannis, Leonzio Rizzod, and Riccardo Secomandi. The ‘Great Lockdown’ and its determinants. Economics Letters, 197:109628, October 2020.

[21] Ibtihal Ferwana and Lav R Varshney. Social capital dimensions are differentially associated with COVID-19 vaccinations, masks, and physical distancing. PLoS ONE, 16(12):e0260818, 2021.

[22] Thom File and Mathew Marlay. Living alone has more impact on mental health of young adults than older adults, January 2021. United States Census Bureau.

[23] James H. Fowler, Seth J. Hill, Remy Levin, and Nick Obradovich. The effect of stay-at-home orders on COVID-19 cases and fatalities in the United States. medRxiv 2020.04.13.20063628., April 2020.

[24] Andrew I. Friedson, Drew McNichols, Joseph J. Sabia, and Dhaval Dave. Did California’s shelter-in-place order work? early coronavirus-related public health effects. Working Paper 26992, National Bureau of Economic Research, April 2020.

[25] Osea Giuntella, Kelly Hyde, Silvia Saccardo, and Sally Sadoff. Lifestyle and mental health disruptions during COVID-19. Proceedings of National Academy of Sciences of the United States of America, 118(9):e2016632118, March 2021.

[26] Andrew Goodman-Bacon and Jan Marcus. Using difference-in-differences to identify causal effects of COVID-19 policies. Survey Research Methods, 14(2):153–158, 2020.

[27] Maria Rosaria Gualano, Giuseppina Lo Moro, Gianluca Voglino, Fabrizio Bert, and Roberta Siliquini. Effects of Covid-19 lockdown on mental health and sleep disturbances in Italy. International Journal of Environmental Research and Public Health, 17(13):4779, June 2020.

[28] Violeta A. Gutkowski. Lockdown Responses to COVID-19. Technical report, Federal Reserve Bank of St. Louis, April 2021.

[29] Xiaolin Huang, Xiaojian Shao, Li Xing, Yushan Hu, Don D Sin, and Xuekui Zhang. The impact of lockdown timing on COVID-19 transmission across US counties. EClinicalMedicine, 38:101035, August 2021.

[30] Luke J. Keele, Dylan S. Small, Jesse Y. Hsu, and Colin B. Fogarty. Patterns of effects and sensitivity analysis for differences-in-differences. 1901.01869 [stat.AP]., January 2019.

[31] Barbara S. Klees, Eric T. Eckstein II, and Catherine A. Curtis. Brief summaries of Medicare & Medicaid. Summary report, Centers for Medicare Medicaid Services, Department of Health and Human Services, November 2020.

[32] Martin Knapp and Gloria Wong. Economics and mental health: The current scenario. World Psychiatry, 19(1):3–14, 2020.

[33] Richard Layard, Andrew Clark, Jan-Emmanuel De Neve, Christian Krekel, Daisy Fancourt, Nancy Hey, and Gus O’Donnell. When to release the lockdown? A wellbeing framework for analysing costs and benefits. IZA Discussion Papers 13186, Institute of Labor Economics (IZA), April 2020.

[34] Minha Lee, Jun Zhao, Qianqian Sun, Yixuan Pan, Weiyi Zhou, Chenfeng Xiong, and Lei Zhang. Human mobility trends during the early stage of the COVID-19 pandemic in the United States. PLoS ONE, 15(11):1–15, November 2020.

[35] Gil Loewenthal, Shiran Abadi, Oren Avram, Keren Halabi, Noa Ecker, Natan Nagar, Itay Mayrose, and Tal Pupko. COVID-19 pandemic-related lockdown: response time is more important than its strictness. EMBO Molecular Medicine, 12:e13171, October 2020.

[36] David McDaid, A-La Park, and Kristian Wahlbeck. The economic case for the prevention of mental illness. Annual Review of Public Health, 40:373–389, 2019.

[37] Weich S. McKenzie K, Whitley R. Social capital and mental health. The British Journal of Psychiatry, 181(4):280–283, October 2002.

[38] Osnat C. Melamed, Margaret K. Hahn, Sri Mahavir Agarwal, Valerie H. Taylor, Benoit H. Mulsant, and Peter Selby. Physical health among people with serious mental illness in the face of COVID-19: Concerns and mitigation strategies. General Hospital Psychiatry, 66:30–33, September-October 2020.

[39] Laura Montenovo, Xuan Jiang, Felipe Lozano Rojas, Ian M. Schmutte, Kosali I. Simon, Bruce A. Weinberg, and Coady Wing. Determinants of Disparities in COVID-19 Job Losses. NBER working paper 27132, 2021.

[40] Geofrey Musinguzi and Benedict Oppong Asamoah. The Science of Social Distancing and Total Lock Down: Does it work? Whom does it benefit? Electronic Journal of General Medicine, 17(6):em230, April 2020.

[41] Sadiq Y. Patel, Ateev Mehrotra, Haiden A. Huskamp, Lori Uscher-Pines, Ishani Ganguli, and Michael L. Barnett. Trends in outpatient care delivery and telemedicine during the COVID-19 pandemic in the US. JAMA Internal Medicine, 181(3):388–391, November 2020.

[42] Betty Pfefferbaum and Carol S. North. Mental health and the Covid-19 pandemic. The New England Journal of Medicine, 383:510–512, August 2020.

[43] David Dooley Prause, JoAnn and Jimi Huh. Income volatility and psychological depression. American Journal of Community Psychology, 43(1):57–70, 2009.

[44] Ashesh Rambachan and Jonathan Roth. An honest approach to parallel trends, November 2020. un-published.

[45] Charles Roehrig. Mental disorders top the list of the most costly conditions in the United States: $201 billion. Health Affairs, 35(6):1130–1135, 2016.

[46] Manuel Serrano-Alarcón, Alexander Kentikelenis, Martin Mckee, and David Stuckler. Impact of covid-19 lockdowns on mental health: Evidence from a quasi-natural experiment in England and Scotland. Health Economics, 31(2):284–296, 2022.

[47] Naomi M Simon, Glenn N Saxe, and Charles R Marmar. Mental health disorders related to COVID-19–related deaths. JAMA, 324(15):1493–1494, 2020.

[48] Linda Thunström, Stephen C. Newbold, David Finnoff, Madison Ashworth, and Jason F. Shogren. The benefits and costs of using social distancing to flatten the curve for COVID-19. Journal of Benefit-Cost Analysis, 11(2):179–195, Summer 2020.

[49] The New York Times. Coronavirus (Covid-19) data in the United States, 2021. dataset.

[50] Jeff Trinkl and Alejandro Muñoz del Río. Effect of COVID-19 pandemic on visit patterns for anxiety and depression. Epic Health Research Network, August 2020.

[51] Marielle Wathelet, Stéphane Duhem, Guillaume Vaiva, Thierry Baubet, Enguerrand Habran, Emilie Veerapa, Christophe Debien, Sylvie Molenda, Mathilde Horn, Pierre Grandgenèvre, Charles-Edouard Notredame, and Fabien D’Hondt. Factors associated with mental health disorders among university students in France confined during the COVID-19 pandemic. JAMA Network Open, 3(10):e2025591, October 2020.

[52] Bret Zeldow and Laura A. Hatfield. Confounding and regression adjustment in difference-in-differences. 1911.12185 [stat.AP]., November 2019.

